# ELISA–on-Chip: High throughput antibody profiling using antigen microarrays

**DOI:** 10.1101/2022.07.05.22277251

**Authors:** Shlomia Levy, Marwa Abd Alhadi, Asaf Azulay, Amit Kahana, Nir Bujanover, Roi Gazit, Maureen A. McGargill, Lilach M. Friedman, Tomer Hertz

## Abstract

Vaccination and natural infection both elicit potent humoral responses that provide protection from subsequent infections. The immune-history of an individual following such exposures is in part encoded by antibodies. While there are multiple immunoassays for measuring antibody responses, the majority of these methods measure responses to a single antigen. A commonly used method for measuring antibody responses is the enzyme-linked immunosorbent assay (ELISA) assay - a semi-quantitative assay that is simple to perform in research and clinical settings. Here we present the ELISA-on-Chip assay - a novel antigen microarray based assay for rapid high-throughput antibody profiling. The assay can be used for profiling IgG, IgA and IgM responses to multiple antigens simultaneously, requiring minimal amounts of sample and antigens. Using three different types of influenza antigen microarrays, we demonstrated the specificity and sensitivity of our novel assay and compared it to the traditional ELISA assay, using samples from mice, chickens and humans. We also showed that our assay can be readily used with dried blood spots, which can be collected from wild birds, as well as from newborns and children. The ELISA-on-Chip assay can be readily used to profile hundreds of samples against dozens of antigens in a single day, and therefore offers an attractive alternative to the traditional ELISA assay.

## Introduction

Immunoassays are a broad set of methods that can be used to detect the presence of immune responses to pathogens, autoantigens or other antigens. Antibodies encode the immune-history of an individual following exposures to both infections and vaccines, and also offer protection from subsequent infections. A common method that has been widely used for antibody characterization is the enzyme-linked immunosorbent assay (ELISA) [1], [2]. The ELISA method can detect the presence of hormones, peptides, proteins and antibodies against a specific antigen of interest. The assay uses an enzyme-substrate reaction that can be measured and quantified by optical density. There are several common ELISA techniques, including direct, indirect, sandwich and competitive ELISAs [3]. The ELISA is a high accuracy semi-quantitative assay that is simple to perform in the lab using a variety of sample types and available reagents and equipment [4]. The ELISA assay is widely used to (1) detect ligands in various biological samples including swabs, blood, sera, and stool; (2) for clinical diagnosis of diseases such as HIV and malaria [5]; (3) and for characterizing antibody responses to vaccines and natural infections [6]. Each ELISA assay can quantify antibodies to a single antigen. Therefore, characterizing binding of a single sample to multiple antigens using this assay is laborious and requires high volumes of sample, and antigen.

Antigen microarrays (AM) are a high throughput antibody binding assay that allows the quantification of antibody responses to hundreds or thousands of antigens simultaneously [PMID: 20450869]. In essence, the AM provides an efficient and highly sensitive antibody binding screen. This platform can accommodate a variety of different antigens, including proteins, peptides, lipids, and whole viruses, providing a comprehensive binding antibody profile. It has been extensively used to study antibody responses to both viral and bacterial infections [7-9], as well as to identify cancer [10] and autoimmune biomarkers [11, 12].

Here we developed and optimized an antigen microarray based assay for rapid high-throughput antibody profiling. We developed an influenza specific AM spotted with recombinant Hemagglutinin (HA) and Neuraminidase (NA) proteins of multiple influenza strains from the H1N1, H3N2, and B subtypes. We demonstrate the specificity of our assay using subtype-specific and cross-reactive influenza monoclonal antibodies. We then characterize the immune-history to previous influenza infections using serum samples from mice and humans. We then demonstrate that the assay can also be used to profile antibodies from dried blood spots, which enables profiling field samples from wild birds. Finally, we develop the ‘ELISA-on-Chip’ assay - an ELISA like antibody binding assay based on antigen microarrays in which each antigen is spotted in several serial concentrations. The ELISA-on-Chip assay allows rapid semi-quantitative characterization of binding profiles to a large panel of antigens simultaneously, using minimal sample and antigen volumes. Using human serum samples, we compare our novel assay to the traditional ELISA assay, and demonstrate its concordance.

## Methods

### Influenza specific antigen microarrays

For a complete list of antigens see **Supplementary Table S1**. We used three types of influenza AMs in this study:

#### 1. Human influenza Ams

Single-concentration influenza protein microarrays which included 28, 46 or 28 (‘hmAbs’, ‘M1’ and ‘M2’ AMs, respectively, in **Supplementary Table 1**) recombinant influenza hemagglutinin (rHA) proteins or rHA1 subunits from human influenza strains. All proteins were spotted at a single concentration (32.5 μg/ml) for screening anti-influenza human monoclonal antibodies (hmAbs), and for profiling serum samples from mice that were exposed to sublethal doses of influenza viruses.

#### 2. Pan-influenza AMs

arrays which included 46 recombinant HA proteins and 14 recombinant NA proteins from human and avian influenza A subtypes and influenza B strains. The arrays also included 4 influenza internal proteins from the PR8 strain: M1, NS1, NS2 and NP. All antigens were spotted as a single-concentration (32.5 ug/ml). Antigens were purchased from Sino Biologics, Native Antigen, or were obtained as a gift from the International Reagent Resource, as described in **Supplementary Table 1**. These arrays (termed ‘C’ in **Supplementary Table 1**) were used for profiling chicken IgY anti-influenza antibodies.

#### 3. ELISA-on-Chip influenza AMs

To compare the ELISA-on-Chip antigen microarray (AM) to the standard ELISA assay, we used rHA proteins from 4 influenza strains that were spotted in 11 serial dilutions. These included three seasonal vaccine strains (north hemisphere): H3N2 A/Wisconsin/67/2005; H3N2 A/Brisbane/10/2007 and H1N1 A/California/07/2009, and the avian influenza H7N9 A/Shanghai/1/2013 strain (EoC AMs in **Supplementary Table S1**).,. All proteins were synthesized by Sino Biological (Beijing, China).

### Mouse serum samples

Nine C57BL/6 mice were infected intranasally (i.n.) with a sublethal dose of A/PuertoRico/8/1934 (PR8) H1N1 influenza virus, as previously described [13], and serum samples were collected pre-infection and 28 days post infection. Another set of 8-week-old C57BL/6 female mice were injected intramuscularly (i.m.) with either one of three viruses; A/X31/1968 (X31, H3N2), mouse-adapted A/California/07/2009 (Cal09, H1N1) or A/Vietnam/1203/04 (Viet1203, H5N1) viruses or none. Each infection group included 10 mice. Serum samples were collected 28 days post infection. The mice experiments were approved by the Ben Gurion University Committee for the Ethical Care and Use of Animals in Experiments, and the Animal Ethics Committee at St. Jude Children’s Research Hospital, respectively.

### Chicken serum and blood drop samples

Blood samples were obtained from 36 breeder chickens that were vaccinated twice with the avian H9N2 2018 influenza vaccine (batch 947, Phibro Israel). Blood samples were collected from 40-41 days old female chicks, 34 days following the first vaccination (n=16); and from 2.5-3 months male and female chickens 30 days following the second dose (n=20). Four blood drops from each sample were dropped on Whatman FTA blood cards (125-500 μl drops), and the rest of the sample was centrifuged for serum isolation. The dry blood drops and sera were stored frozen at -20oC. For microarray experiments, each blood drop was incubated in 2 ml 0.05% PBST (0.05% tween-20 in PBS) overnight on a shaker, and the liquid was collected and stored frozen.

### Human monoclonal antibodies

A set of 8 human monoclonal antibodies (hmAbs) isolated from subjects vaccinated with various influenza vaccines [14, 15] are described in Supp. Table 2.

### Human serum samples

Serum samples were collected from 10 healthy young men in Israel in January 2018, from a clinical study approved by the Institutional Review Board of the Israeli Defense Force (approval number 1854-2017, IDF IRB).

### ELISA assay

To run efficient ELISA assays with reduced sample volumes and antigens, we optimized our ELISA assay for 384 well plate format, using a liquid dispensing robot (EzMate 601, Arise Biotech Corp.). 384 well white Maxisorp-coated plates (120μl wells, cat# 460372, ThermoFisher, USA) were coated with 17 μl of 4 μg/ml recombinant hemagglutinin (rHA) protein per well (diluted in PBS) and incubated overnight at 4°C. Plates were washed 5 times with PBS-T washing buffer (0.1% Tween-20 in PBS, 60 μl per well) using a plate-washer (ELx405™ Select Deep-Well Microplate Washer, BioTek™). Plates were then blocked with 100 μl of 10% skim milk powder (Sigma) in PBS-T and incubated for 1 h at 37°C. Following 5 PBS-T washes, human serum samples were diluted in 2-fold serial dilutions (1:25 - 1:409600) in 2% skim milk in PBS-T, and added to the plates in triplicates (30 μl per well) for 1 h incubation at 37°C in the incubator, and washed. The secondary antibody, Peroxidase-AffiniPure Goat Anti-Human IgG (H+L) (cat# 109-035-088, Jackson) and Anti-Mouse IgG (H+L), HRP Conjugate, (cat# W4021, Promega), was diluted 1:10,000 and 1:2500 respectively in 2% skim milk in PBS-T and added to the plates (30 μl per well) following 5 PBS-T washes. After incubation for 1 h at 37°C and washes, equal volumes of Peroxide and Luminol were mixed and added (60 μl per well, SuperSignal West Pico Chemiluminescent Substrate, cat# 34579, ThermoFisher). Following 1 min incubation, the luminescence was measured by an ELISA reader (Infinite^®^ 200 PRO, TECAN) at 600 nm wavelength.

### ELISA analysis

Median OD values of each triplicate were calculated and negative control of 2% skim milk was subtracted to get the relative light units (RLU). ELISA curves were fitted using a 5 parameter logistic model:

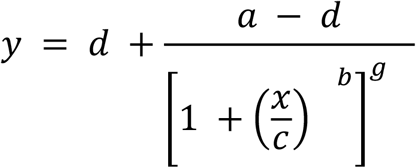

Where *a* is the minimal value obtained; *b* is the slope of the curve; *c*, is the inflection point; *d* is the maximal value; and *g* is the asymmetric factor. The model was fitted using the curve fit function of the scipy.optimize library in Python.

### Antigen microarray spotting

Recombinant influenza proteins were spotted onto *N*-hydroxysuccinimide ester–derivatized Hydrogel slides (H slides) using a Scienion Sx non-contact array spotter. For ELISA-on-Chip AMs four rHA influenza proteins were spotted at 11 serial concentrations in the range of 125 μg/ml - 122 ng/ml in Scispot D1 spotting buffer (Scienion, Germany). Recombinant proteins for the pan-influenza and human-influenza protein microarrays were spotted at a single concentration of 32.5 μg/ml in the Scispot D1 spotting buffer (mice and human serum samples), or at a single concentration of 16.25 μg/ml in 0.01% triton X-100 (chicken experiment). Three different types of broader influenza microarrays were spotted as described above (see **Supplementary Table 1**). Spot volumes ranged between 300-360pL. Antigens at each concentration were spotted in triplicates. Sixteen identical microarrays were spotted on each microarray slide. Each printing batch included up to 140 microarray slides, yielding a total of up to 2,240 arrays per batch.

### Antigen microarray assay

Array slides were blocked with 4 ml per slide of chemical blocking solution (50 mM ethanolamine, 50 mM borate, pH 9.0) for 1 h at room temperature (RT) on a shaker. After blocking the liquid was vacuumed, the slide was washed 2 times for 3 minutes in a washing buffer (0.05% tween-20 in PBS), 2 times for 3 minutes in PBS and an additional 3 minutes wash in double deionized water (DDW). Every wash was performed with 3ml of liquid per slide on a shaker at RT. Samples were diluted in a hybridization buffer (1% BSA / 0.025% tween-20 in PBS). Human serum samples were diluted 1:1000, mice serum samples were diluted 1:100, chicken serum samples were diluted 1:4000, and chicken dry blood spots (∼500 μl) were reconstituted in 2 ml washing buffer were diluted 1:20. Human mAbs were incubated in three serial concentrations: 6, 1.5 and 0.375 μg/ml.

Following 2 hours incubation, the slides were dried by centrifugation at RT for 5 minutes at speed 2000 rpm in a slide holder padded with kim wipes, loaded on divided incubation trays (PepperChips, PepperPrint, Germany), and then the samples were added and hybridized with the arrays for 2h at RT on shaker. After hybridization, the samples were discarded and the slides were washed in washing buffer X 2 and PBS X 2 as described above. After washes, the slides were incubated for 45 minutes on the shaker at RT with a fluorescently labeled polyclonal secondary antibody in the hybridization buffer. The secondary antibody for human serum sample and human mAbs was Alexa Fluor® 647 affinipure Donkey Anti-Human IgG (H+L), (cat# 709-605-149, Jackson ImmunoResearch) used at 1:1000 dilution. The secondary antibody for mouse serum samples was Alexa Fluor® 647 conjugated AffiniPure Goat Anti-Mouse IgG Fcγ Fragment Specific (cat# 115-605-008, Jackson ImmunoResearch), used at a 1:3000 dilution (Fig. 1) or 1:4000 dilution (Fig. 2). The secondary antibody for chicken serum and dry blood drop samples was Alexa Fluor® 647 AffiniPure Goat Anti-Chicken IgY (IgG) (H+L) (cat# 103-605-155, Jackson ImmunoResearch) used at a dilution of 1:1000 for serum samples and at a dilution of 1:2000 dilution for dry blood spot samples. To detect bound immunoglobulins, slides were scanned on a three-laser GenePix 4400 scanner. Images were analyzed using GenePix Pro version 7 to obtain the mean fluorescence intensity (MFI) of each spot after subtracting the mean local background fluorescence intensity (0 ≤MFI ≤ 65,000).

**Figure 1:**
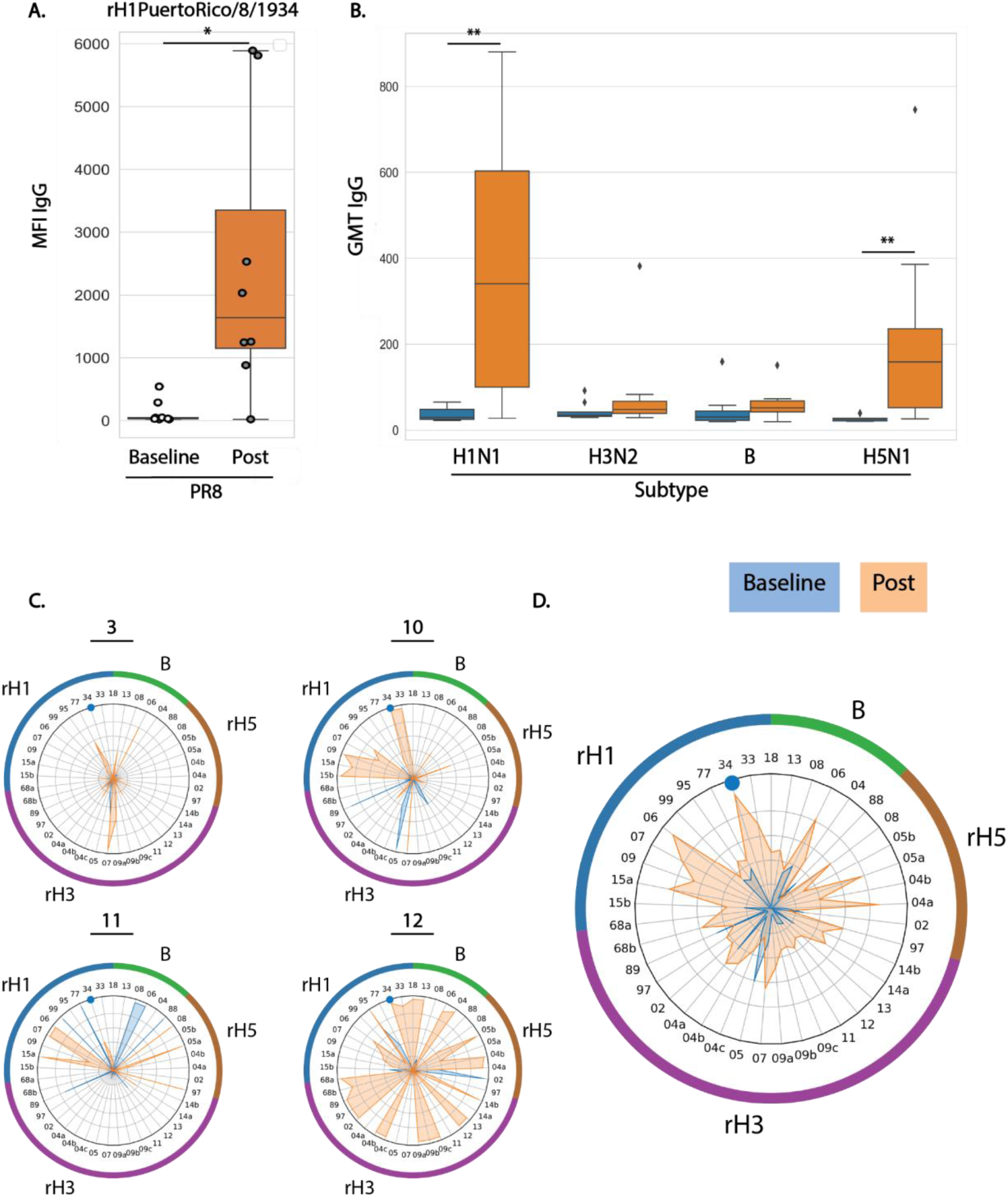
Magnitude, breadth and specificity of anti-influenza HA IgG response to A/PuertoRico/8/1934 (PR8) sublethal infection in mice, as measured by antigen microarrays spotted with recombinant HA (rH) proteins from 41 influenza strains. Nine C57BL/6 mice were infected intranasally (i.n.) with A/PuertoRico/8/1934 (PR8) H1N1 viruses, as previously described [13]. Serum samples were collected pre-infection (blue) and 28 days post infection (orange). Antigen microarrays spotted with 41 recombinant full HA proteins or HA1 units of 36 influenza A strains from 3 subtypes (11 rH1 proteins from H1N1 strains, including the PR8 infection strain; 18 rH3 proteins from H3N2 strains; and 7 rH5 proteins from H5N1 strains), and 5 influenza B strains, were used to measure IgG binding to each spotted protein by mean fluorescence intensity (MFI). **(A)** IgG binding to the rHA protein of the infection strain PR8 (MFI). **(B)** Geometric mean magnitudes (GMT) of IgG MFI of all the spotted proteins from each subtype. In panels A and B horizontal lines represent the median, boxes denote the 25th and 75th percentiles, and the error bars represent 1.5 times the interquartile range. Statistical significance was assessed using the Wilcoxon signed rank test (pre vs. post): * p<0.05., ** p<0.005. **(C)** Spider plots of individual IgG profiles of binding to the microarray proteins in baseline (blue) and post-PR8 infection (orange) serum samples from 4 representative mice. The ID number of each mouse is presented above each spider graph. **(D)** A spider plot of the mean IgG binding profiles across all the 9 mice at the two timepoints. In panels C and D, each vertex represents the normalized MFI to a single rHA protein. The numbers listed around the inner circle denote the year each influenza strain was isolated. Counter-clockwise, rHA proteins from: H5N1 strains 1997-2008 (brown); H3N2 strains 1968-2014 (purple); H1N1 strains 1918-2015 (blue); and B strains 1988-2013 (green). The blue dot labels the infection strain PR8. See Supplementary Table 1 for a list of strain names.

**Figure 2:**
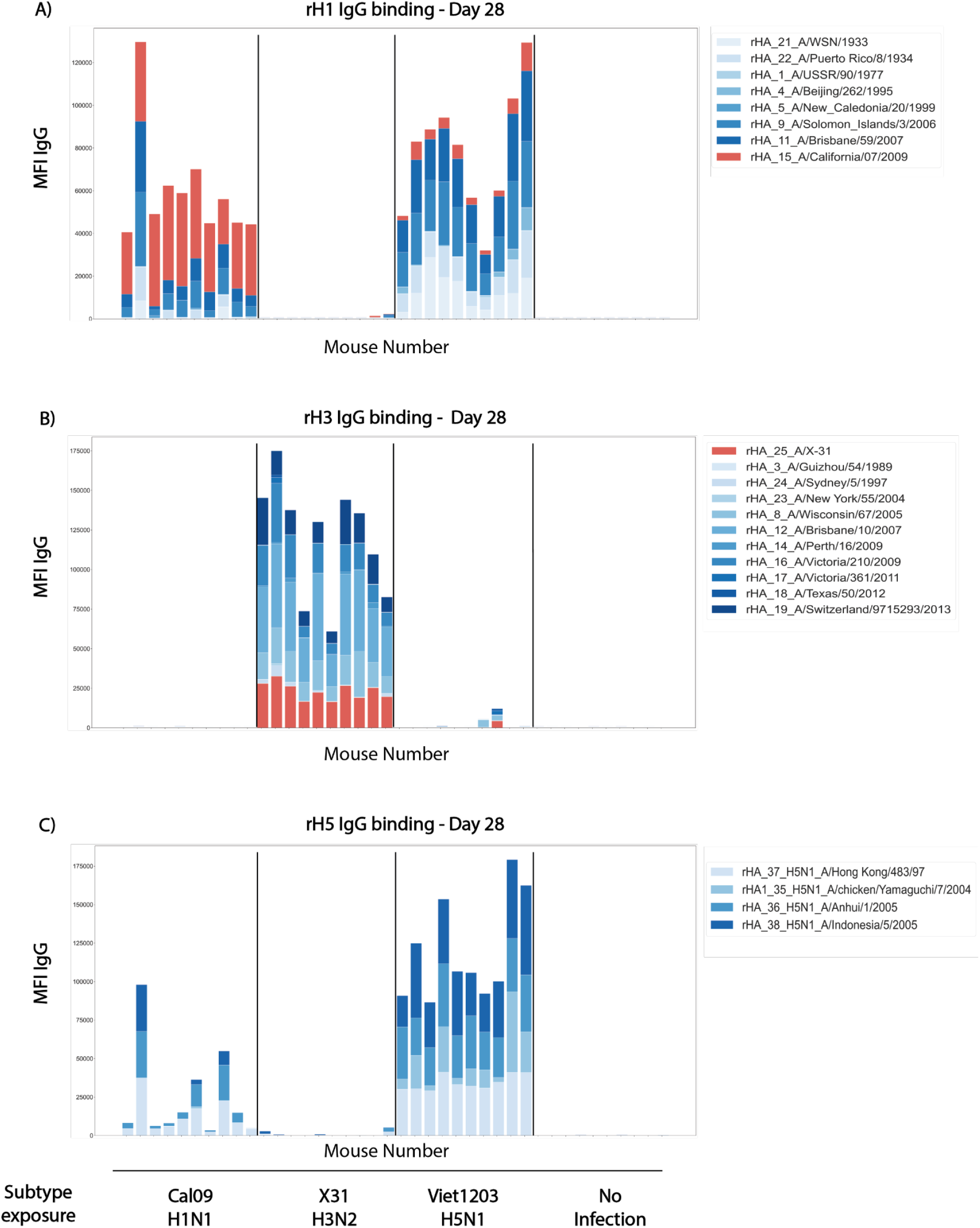
Subtype specificity of influenza rHA microarrays. Four groups of mice (n=10 per group) were injected intramuscularly (IM) with sublethal doses of influenza A viruses from 3 different subtypes: pandemic A/California/07/2009 (Cal09, H1N1), A/HKx31 (X31, H3N2) and A/Vietnam/1203/2004 (Viet1203, H5N1). A negative control group was not exposed to any virus. Serum samples were collected at day 28 post infection and binding of IgG antibodies to influenza recombinant hemagglutinin (rH) proteins were profiled using an influenza antigen microarray (microarray M2 in supplementary Table 1). **(A)** IgG responses to H1N1 strains (rH1 n=8), **(B)** IgG responses to H3N2 strains (rH3, n=11), and **(C)** IgG responses to H5N1 strains (rH5, n=4). Each bar represents the cumulative MFI for a single mouse to all the rH proteins listed. Response to the vaccine strain is in orange.

### Antigen microarray analysis

The array results were analyzed using an in-house pipeline developed in python. Since each antigen at each concentration was spotted in triplicate, the median MFI intensity of each triplicate was calculated. During each experiment, a negative control array was hybridized with the hybridization buffer only. The background staining of the negative control array was subtracted from all other arrays. For antigens that were spotted in serial concentrations, a 5-parameter logistic regression model was used to fit curves to the measured median fluorescence intensity (MFI) vs the antigen concentration, and the area under the curve (AUC) was calculated. For the influenza microarrays, we divided the rHA and rNA proteins into groups according to their subtype. The magnitude of antibody response to a group of antigens was defined as the sum of MFI levels to all the proteins included in the group. To compare groups with different numbers of proteins, the geometric mean magnitude was calculated.

### Statistical analysis

Comparisons between experimental groups, and between the traditional ELISA and ELISA-on-Chip assays were performed using the Wilcoxon ranksum test. Correlations were computed using Pearson correlation, or when relevant Spearman correlation. All analysis was performed in Python.

## Results

### Monovalent influenza infection in mice generates a subtype specific antibody profile

To test the specificity and sensitivity of the antigen microarrays, mice (n=10) were infected intranasally (i.n.) with a sublethal dose of A/PuertoRico/8/1934 (PR8) (A/H1N1). Serum samples were collected at baseline and 28 days post infection. IgG antibody profiles from each mouse were generated at baseline and day 28, using the Human-influenza AMs spotted with recombinant HA proteins from 11 A/H1N1 (rH1), 18 A/H3N2 (rH3), 7 A/H5N1 (rH5) and 5 influenza B strains (see methods). At baseline, mice displayed none, or very low levels of IgG binding to the PR8 rHA protein. Post-infection anti-PR8 antibody levels were significantly higher than baseline levels (p = 0.007, Figure 1A). The anti-PR8 antibody responses varied significantly between mice (Figure 1A) and one mouse failed to mount an immune response following sub-lethal infection. Geometric mean magnitude titers (GMT) were computed for antibody reactivity to each of the four influenza subtypes. Baseline titers were low to all four subtypes, while only H1 and H5 titers significantly increased after PR8 infection (p=0.003 and p=0.002 respectively, Figure 1B). Previous work reported that antibodies developed following H1N1 infection or vaccination can cross-react with HA proteins of H5N1 strains (e.g. [16-20]).

To visualize the baseline and post vaccination IgG profile of each individual mouse separately, we generated spider plots to display the normalized MFI staining for all rH1, rH3, rH5 and B rHA antigens. Representative spider plots for four mice are presented in **Figure 1C**. Most mice generated very weak baseline responses to the rHA proteins. Each mouse has a unique IgG profile that varies both in the overall breadth and magnitude of responses, but also in the specificity to each subtype. For example, mouse number 12 had a very broad post-infection response, including high IgG binding to all the three influenza A subtypes, as well as to influenza B rHA proteins. On the other hand, mouse number 10 responded predominantly to rH1. We computed the average IgG binding of all the 10 mice to each rHA protein at both time points (**Figure 1D**). We found that the highest IgG post-infection responses were against the infection strain (PR8, **Figure 1D**) and the H1N1 A/SolomonIslands/3/2006 strain. However, post-infection IgG levels were also observed for strains from all other influenza A subtypes.

To further examine the specificity of the antigen microarrays, we analyzed serum from mice exposed to different influenza A subtypes. C57BL/6 mice were given either A/HKx31 (X31, subtype H3N2), mouse-adapted A/California/07/2009 (Cal09, subtype H1N1), or A/Vietnam/1203/04 (Viet1203, subtype H5N1), intramuscularly. Serum samples were collected 28 days later, and were incubated with antigen microarrays spotted with 8 rH1, 11 rH3 and 4 rH5 proteins. Mice given Cal09 generated an IgG response predominantly to rHAs from H1N1 subtypes (**Figure 2A**). All the mice that were infected with Cal09 generated high IgG responses to both rH1. Moreover, a high IgG binding to the Cal09 rHA protein was measured. All mice from the Cal09 group generated a significant response to the closely related A/Michigan/45/2015 strain that has 97% identity with Cal09 (**Figure 2A**). High responses to H1N1 viruses before the 2009 pandemic were observed only for a single Cal09 injected mouse, which also developed a significant cross-reactive IgG response to H5N1 strains (compare **Figures 2A** and **2C**). Significant levels of anti-rH3 antibodies were detected only in mice that were injected with the A/H3N2 X31 strain. Weak cross-reactive IgG binding to rH3 proteins were observed in two mice that were injected with the H5N1 strains (**Figure 2B**). The X31 injected mice generated none or very weak antibody responses to rHA proteins from the H1N1 and H5N1 subtypes. All the mice that were infected with the H5N1 Viet1203 strain developed high levels of IgG antibodies to both rH1 and rH5 proteins, which belong to the same antigenic group (**Figure 2C**)[21, 22]. Thus, the antigen microarrays reflect the dominate antibody response to the subtype the mice were exposed to, and detect cross-reactive antibodies specific for multiple subtypes.

### Antibody profiles generated from dry blood spots are comparable to serum antibody profiles following H9N2 influenza vaccination in chickens

Collecting dry blood drops provides an attractive alternative to serum collection since it does not require centrifugation and freezing, and is also minimally invasive, allowing collection of samples in field studies of wild birds, and from newborn babies. To compare the anti-influenza antibody repertoires in serum and dry blood spots that were produced from the same animal, we used blood samples collected from 36 breeder chickens that were vaccinated twice with an H9N2 influenza vaccine. Blood samples were collected from 40-41 days old chicks, 34 days following the first vaccination (n=16), and from 2.5-3 months old chickens a month post the second vaccination (n=20). Four blood drops (125-500 μl) from each sample were dropped on a Whatman FTA blood card and left to dry, while the rest of the sample was centrifuged for serum isolation. IgY antibodies are the major antibody isotype in birds, similar to the IgG isotype in mammals. To compare the binding profiles of IgY from dry blood drops and sera to influenza antigens, we used the pan-influenza AMs (see methods) spotted with 64 recombinant influenza proteins from both human and avian strains and 4 PR8 internal proteins: M1, NS1, NS2 and NP which are relatively conserved. Since only a small volume of serum could be collected, in particular from chicks, samples were run at a dilution of 1:4000. In contrast, each dry blood drop was reconstituted in 2ml buffer and further diluted only by 1:20 for microarray incubation. As a result, some antibody responses were observed only using the dry blood spots (**Figure 3A**).

**Figure 3:**
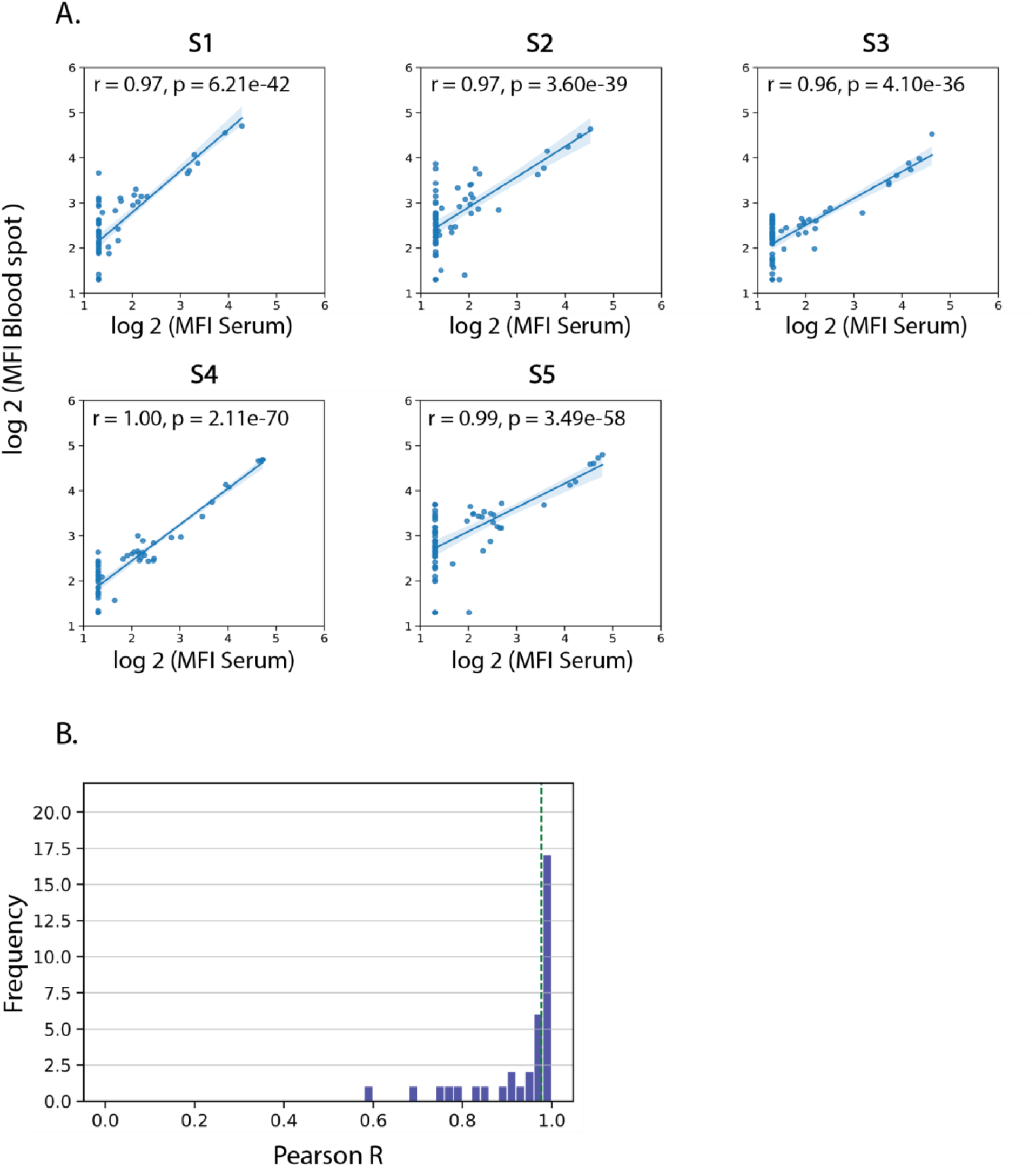
IgY Antibody profiles of dried blood spots are highly correlated to those measured from serum. Chickens were vaccinated with an H9N2 influenza vaccine twice, and blood samples were collected 34 days after the first vaccination (n=16, e.g. samples s1 and s2 are from 41 days old female chicks) or 30 days following the second vaccination (n=20, e.g. samples s3 and s4 from 2.5 months old male chickens, and sample s5 from a 3 months old female chicken). From each blood sample both serum and dry blood spots were collected. The IgY response to vaccinations was profiled using an antigen microarray spotted with recombinant influenza proteins from 15 influenza A and B subtypes (see Supp. Table 1).**(A)** Scatter plots of the microarray MFI results for each serum-blood spot pair of the same subject are presented for 5 samples. The Pearson correlation was computed for each sample. **(B)** The distribution of Pearson correlation values of the microarray results of the 36 serum-blood spot pairs. The dotted green line represents the median r value = 0.977. The median p value was 7.00e-43.

Nevertheless, we found high correlations between IgY levels measured from serum and dry blood drops with an average correlation of 0.932 (p < 2e-7, Pearson correlation, **Figure 3A-B**). Only 2 samples had Serum-blood drop correlations below 0.9 in 2.5-3 months old chickens. Similarly, correlations were lower in 6/16 40-41 days old chicks. In all of these samples there were IgY responses that were detected in the dry blood spots but not in sera (**Supp. Figure 1**). We also observed no background non-specific binding to our arrays when using dry blood spots. This is in contrast to serum samples from some of the chickens that had significant levels of non-specific binding (data not shown).

### Rapid profiling of a panel of human monoclonal antibodies using the ELISA-on-Chip assay

Recent advances in the ability to isolate human monoclonal antibodies (hmAbs) following vaccination or infection [23-25] have highlighted the importance of rapid profiling of their binding specificities. We therefore selected a representative set of 8 human monoclonal antibodies (hmAbs), which were isolated from human individuals that were vaccinated with monovalent or quadrivalent influenza vaccines [14, 15] (summarized in **Supplementary Table 4**). Each of the 8 hmAbs were previously tested against specific influenza antigens to classify their binding to influenza H1N1, H3N2 and (in some cases) B subtypes, and their binding to the HA head or stalk region [14, 15, 26-28]. We profiled each of the 8 hmAbs using the Human-influenza AMs - influenza antigen microarrays that were spotted with a panel of 38 rHA antigens (**Figure 4**). Each protein was spotted in a single concentration of 32.5 μg/ml. All antibodies were profiled in 3 dilutions (6 ug/ml, 1.5 ug/ml, 0.375 ug/ml), and the area under the curve (AUC) was computed for each hmAb across the 3 dilutions. We found that overall, antibodies bound to rHA antigens from the specific subtypes that they were previously reported to bind, i.e hmAb 047-1G05 binds to all tested H1N1 strains exclusively as reported before, while 030-09 2B03 is more specific for pH1N1 strains. However, in some cases, they also bound/not bound influenza antigens from additional subtypes, against which they were not previously tested, i.e 030-09 1E05 hmab was reported to bind influenza B which was not been detected here, and FI6 hmab who was not reported to bind influenza B did exhibit this binding.

**Figure 4:**
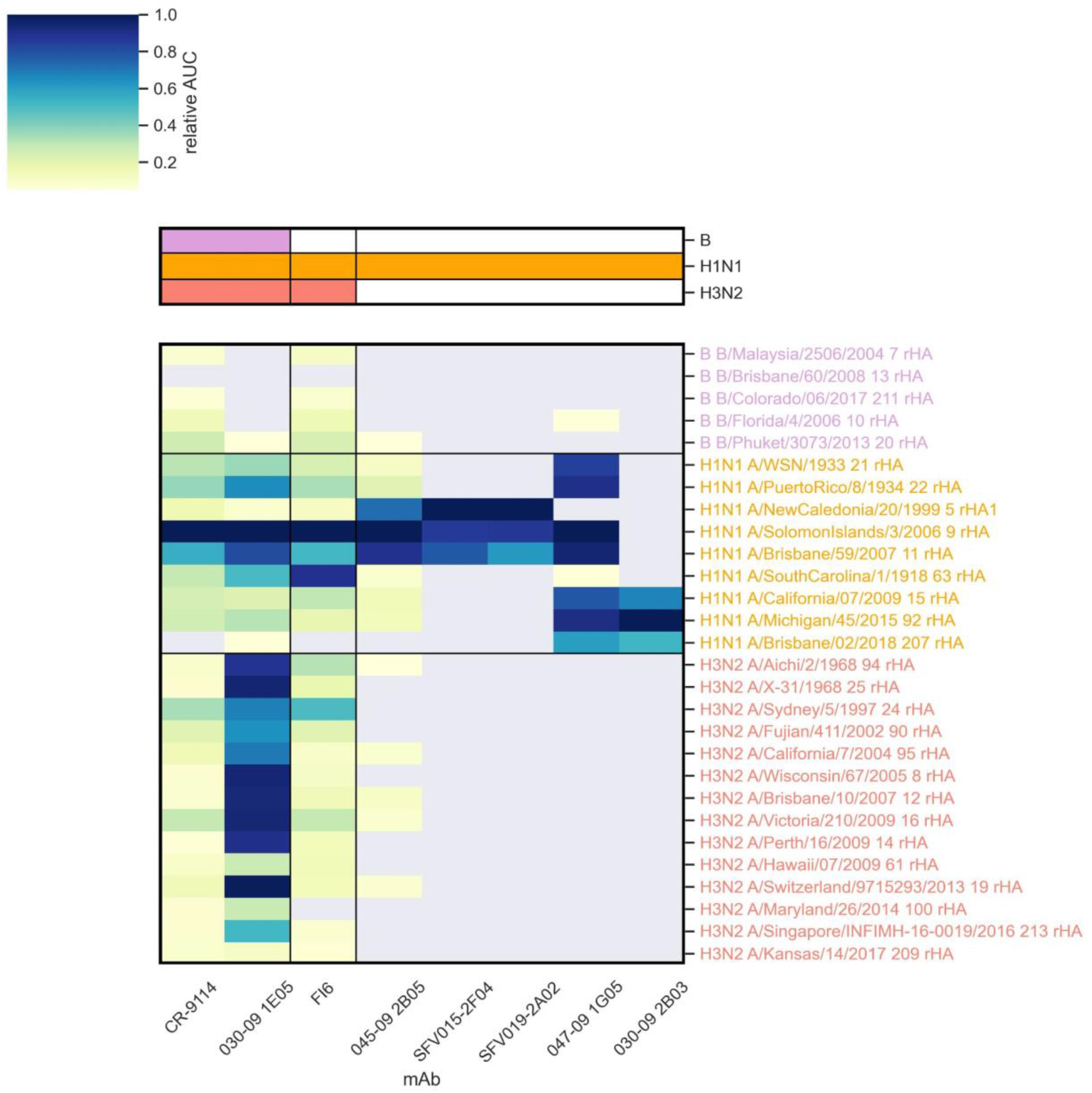
Rapid profiling of a panel of human monoclonal antibodies (hmAbs) using the AM on Chip assay. Relative area under the curve (AUC) results for 8 influenza hmAbs. Each of the hmAb is normalized to its maximal AUC value. AUC were calculated from MFI results of 3 different dilutions from initial concentration (6 ug/ml, 1.5ug/ml, 0.375ug/ml). Each hmAb has previously been tested for binding to specific HA antigen (H1, H3 or B) and for additional properties which are presented on the top bar (see **Supp. Table 2**). AUC values that were smaller than 5% of maximal AUC for each mAb were considered as background (gray). Antibody names in red are HA stalk specific, and those in blue are HA head specific.

### Developing and optimizing an ELISA-on-Chip assay

The accuracy of the traditional ELISA assay derives in part from testing each sample in several serial dilutions, which enables calculation of the area under the curve (AUC) statistic. This approach compensates for cases in which the antigen coating is not in the optimal concentration (see for example **Supplementary Figure 1**, bottom 4 rows). When antigen microarrays are spotted with a single concentration for each antigen, as in the experiments presented above, the incubation of a set of microarrays with a set of serially diluted samples is not efficient, and sometimes impossible when sample volume is limited. We therefore sought to develop the ‘ELISA-on-Chip’ assay - an alternative AM-based assay n which each antigen is spotted in several serial concentrations, such that incubation with a single dilution of the sample can allow computing an area under the curve across dilutions of each of the antigens spotted on the array.

We spotted ELISA-on-Chip microarrays with four influenza rHA proteins. Each protein was spotted in 11 serial two-fold dilutions (5 ng/ml - 125 μg/ml) in triplicates. We used a mouse monoclonal antibody (mAb) to the spotted H3N2 A/Brisbane/10/2007 rHA protein as a primary antibody for assay calibration. The same A/Brisbane/10/2007 rHA protein was also used to coat 384-well ELISA plates at a single concentration (4 μg/ml) to compare the ELISA-on-Chip results to the traditional ELISA assay. Serial concentrations of the primary mAb (5 ng/ml - 80 μg/ml) were hybridized with the ELISA-on-Chip arrays and with the coated ELISA plates. A secondary anti-mouse IgG antibody was used to detect the binding of the primary antibody to the antigen. For the traditional ELISA assay, the secondary antibody was bound to an HRP enzyme, and for ELISA-on-Chip assay the secondary antibody was conjugated to a fluorescent dye (Alexa 635). We used a 5-parameter logistic regression model to fit curves to the measured median fluorescence intensity (MFI) and relative light units (RLU) as a function of the antigen concentration in the ELISA-on-Chip and sample dilution in traditional ELISA, respectively. We then calculated the area under the curves (AUC) for each antigen. We found that the Pearson correlation between the curves of the traditional ELISA and ELISA-on-Chip assays was: *r* = 0.975 (p = *7*.*1* × *10*^−*10*^, Pearson correlation, **Figure 5A**).

**Figure 5:**
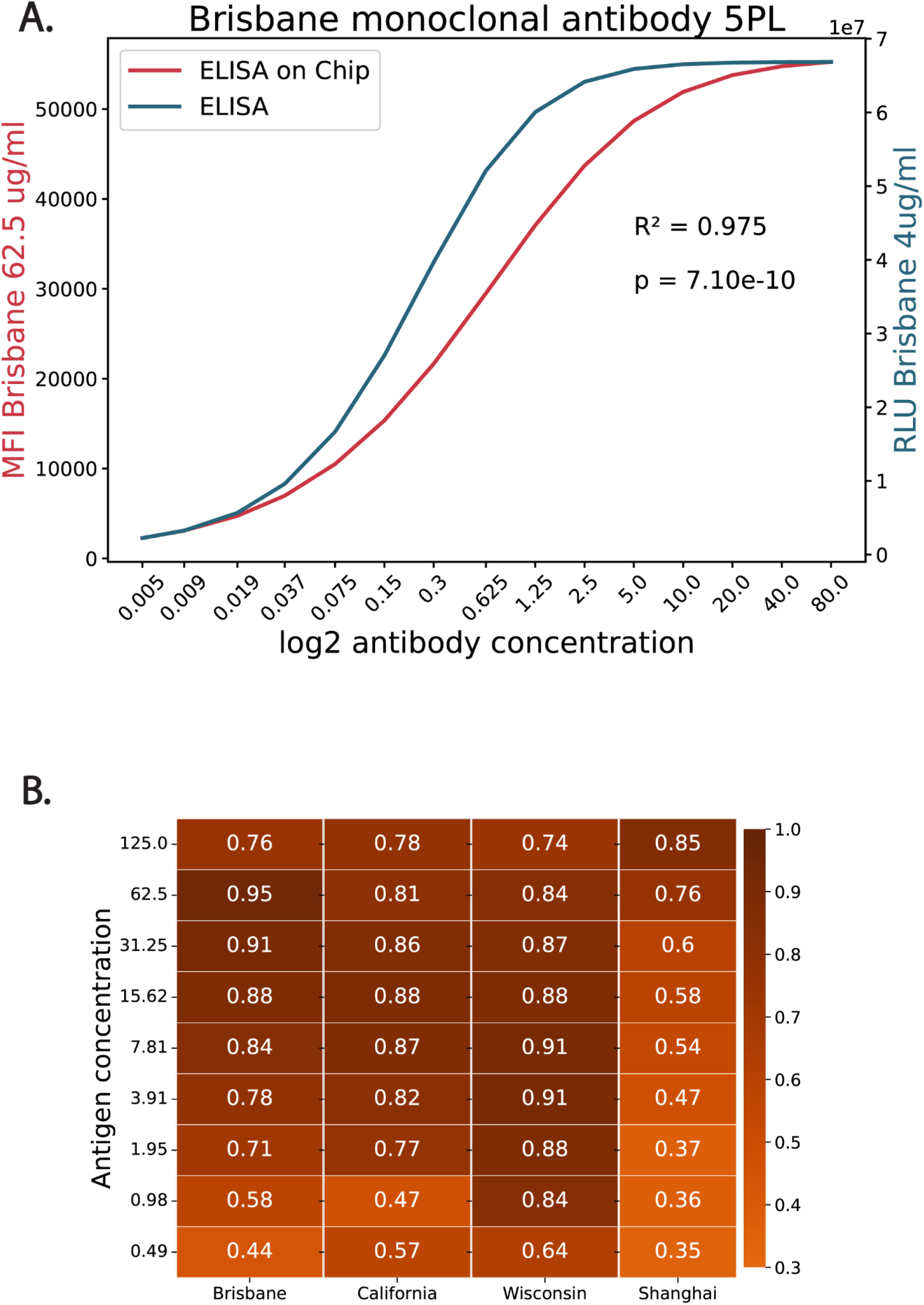
ELISA and ELISA-on-Chip titer curves Pearson correlation. **(A) Monoclonal antibody titer curves:** binding of 15 serial 2-fold dilutions of anti-A/Brisbane/10/2007 rHA monoclonal antibody to the A/Brisbane/10/2007 rHA protein at a single concentration (4 or 62.5 μg/ml), as measured by ELISA (blue) or ELISA on Chip (red), respectively. The curves were fitted using a 5 parameter logistic regression model. The Pearson correlation between the two curves was: r = 0.975, p = 7.1×10^−10^. **(B) Human samples titer curves correlation:** IgG antibodies against 4 Influenza strains were quantified in 10 healthy adult individuals using both ELISA and ELISA-on-Chip assays. ELISA was performed over 15 serial (2-fold) sample dilutions using a single antigen concentration (4 μg/ml). The ELISA-on-Chip was run against antigens spotted in 11 serial dilutions (2-fold, as listed in the Y axis). Pearson correlation coefficients were computed for titer curves for each antigen concentration in ELISA-on-chip compared with the single antigen concentration in ELISA.

We then used serum samples from 10 healthy adults that were collected in a clinical study in Israel during the 2018 winter season. Levels of serum IgG antibodies to rHA proteins of four influenza strains were measured using the traditional ELISA assay and the ELISA-on-Chip assay. The Pearson correlation between the curves generated by the two assays, across all the antigen concentrations spotted on the array, were high for the three human influenza rHA proteins for all relevant concentrations (0.71 <= r <= 0.95, p < 0.0001 **Figure 5B**). However, lower correlations were observed for the rHA of the H7N9 Shanghai strain, an avian strain to which the individuals were not exposed (**Figure 5B**).

### Generating binding curves using antigen dilutions

An important advantage of the antigen microarray assay is that antigens can be spotted in multiple dilutions on each array, allowing us to also characterize antibody binding using a single dilution of the sample, while considering responses to each antigen across all of its dilutions. We used this assay to test the anti H3N2 Brisbane 2007 mouse monoclonal antibody at different concentrations. We found that the assay was able to distinguish between the different concentrations of the same mAb (**Figure 6A**). Since the ELISA-on-Chip AM included the spotted A/Brisbane/10/2007 rHA antigen in serial concentrations and was incubated with serial concentrations of the mAb as in traditional ELISA (ELISA-array assay), we can plot antibody levels levels as a function of both mAb and antigen concentrations. We found that the signal decay across antigen dilutions or mAb dilutions was similar (**Figure 6B**).

**Figure 6:**
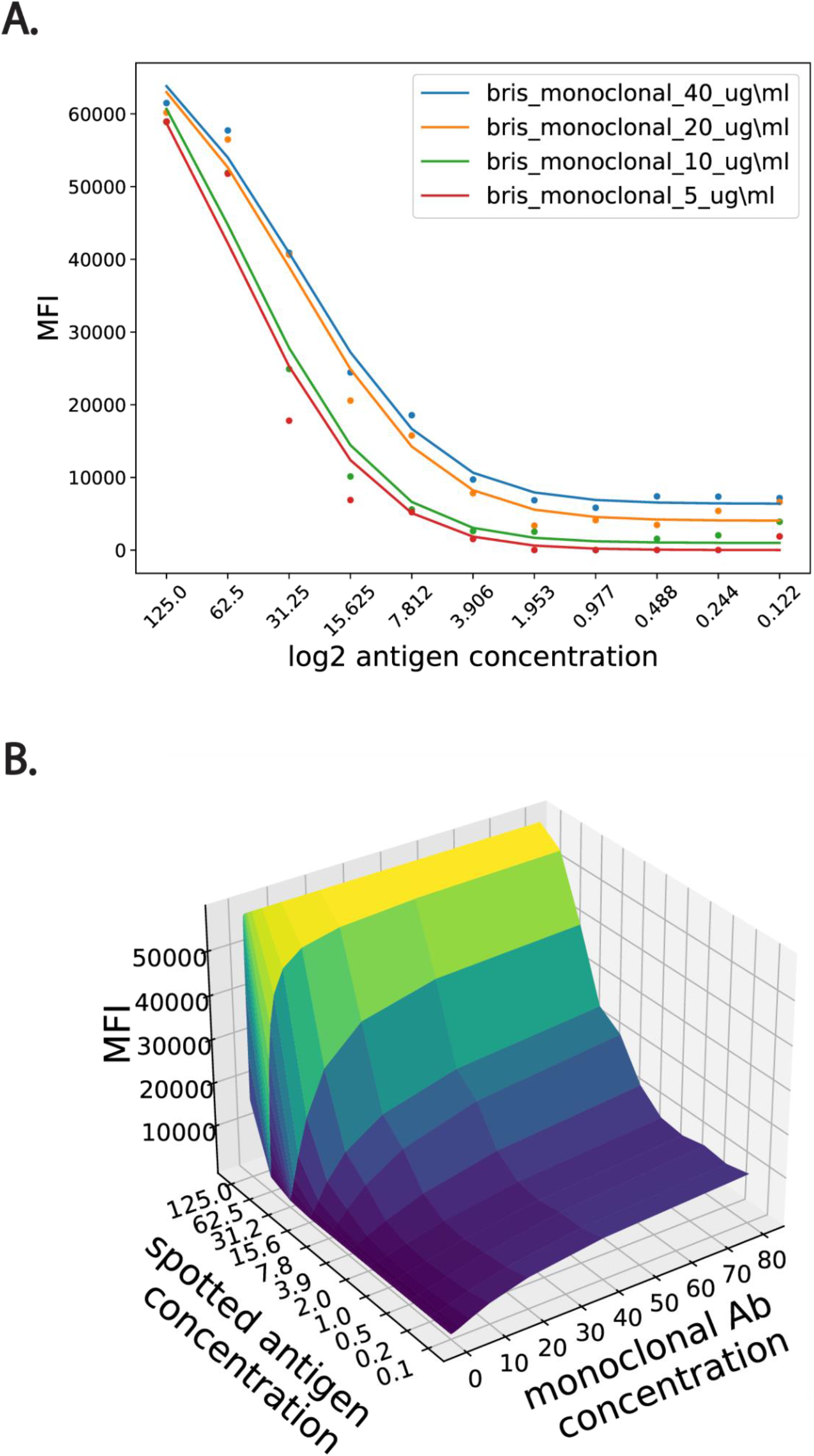
ELISA on Chip antigen dilution titer curves. **(A) Monoclonal antibody (mAb) titer curves:** Example titer curves of the binding level of 4 concentrations of the anti-H3N2 Brisbane 2007 mAb with 11 serial 2-fold dilutions of the spotted A/Brisbane/10/2007 rHA antigen. Each dilution of the mAb was incubated with a different antigen microarray (AM). **(B) Three-dimensional titer curves:** mAb titer curves for antibody serial dilution and spotted antigen serial dilution.

To compare the performance of the traditional ELISA to the ELISA-on-chip assay, we computed the area under the curve (AUC) for each assay dilution curve. Spearman correlation coefficients were computed for AUC in ELISA-array assays that were performed for 10 adult samples diluted 1:3200, and for the traditional ELISA AUC computed across all sample dilutions using a single antigen concentration. Spearman correlation coefficients were: Bris07: *r*_*s*_ = 0.89; Cal09: *r*_*s*_ = 0.92; Wis05: *r*_*s*_ = 0.75; and Shang09: *r*_*s*_ = 0.72. We also computed the pairwise correlations between each ELISA assay for the four different antigens which included 2 H3N2 strains (A/Wisconsin/67/2005, A/Brisbane/10/2007), the H1N1 A/California/07/2009 and the H7N9 A/Shanghai/1/2013. These correlations were also computed for the ELISA-on-Chip assay. We found that overall, there were higher inter-correlations between the ELISA assays, even for strains from different subtypes (**Figure 7, Supplementary Tables 2-3**). These data suggest that the ELISA-on-Chip assay is more subtype specific than the ELISA assay.

**Figure 7:**
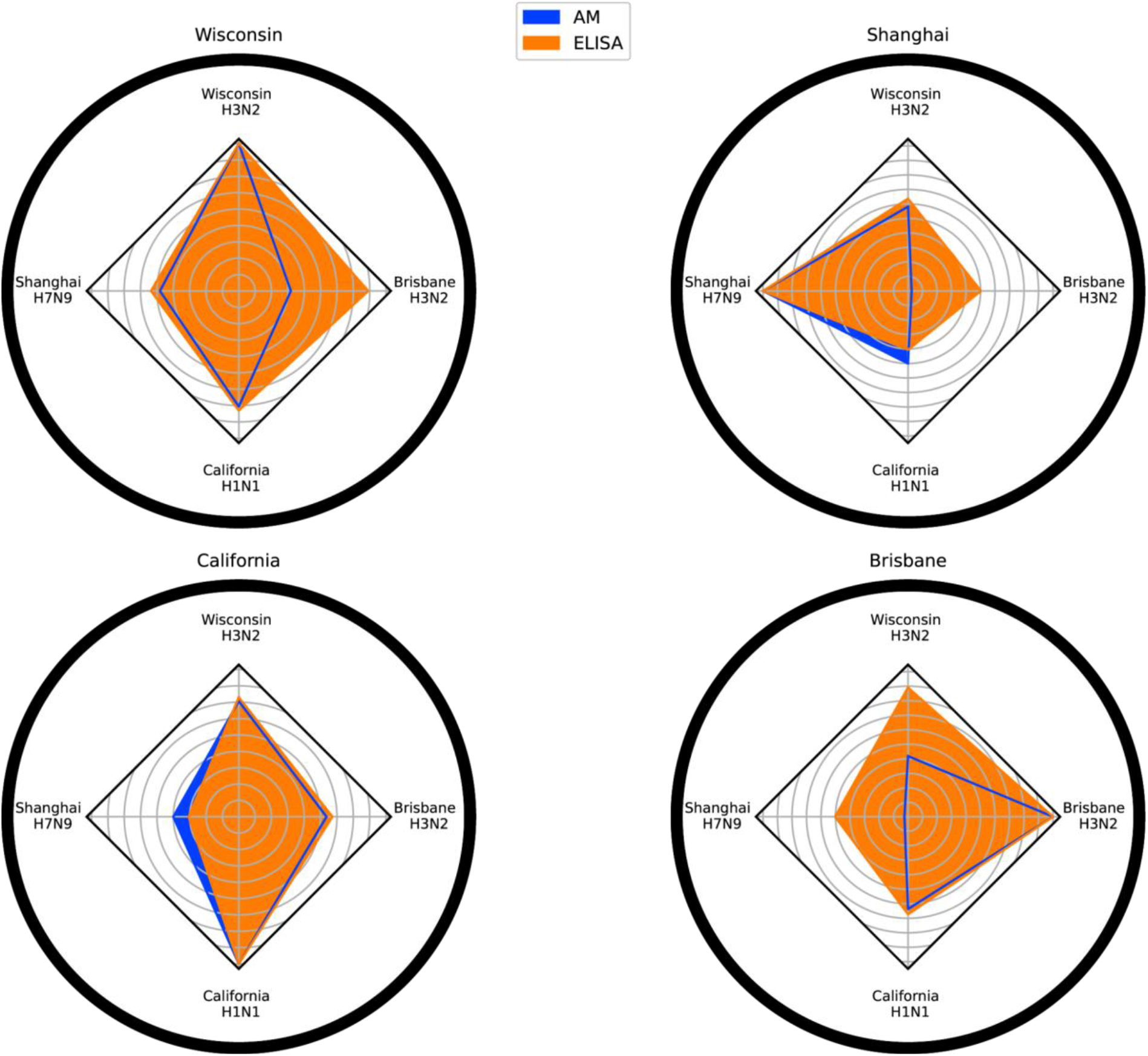
ELISA and AM correlations. We compared the area under the curve of 10 healthy adult individuals using our influenza antigen microarrays (diluted 1:3200, blue) and using traditional ELISA in a single antigen concentration (orange). The Spearman correlation between the AUC scores was computed for all pairwise combinations of four influenza strains: two H3N2 strains A/Wisconsin/67/2005, A/Brisbane/10/2007, the H1N1 A/California/07/2009 and the H7N9 A/Shanghai/1/2013

## Discussion

Here we presented ELISA on a Chip - a novel antigen microarray binding assay that can be used as a high-throughput alternative to the ELISA assay for semi-quantitative profiling of antibody levels to many antigens concurrently. As proof of concept, we used several types of influenza antigen microarrays spotted with recombinant HA proteins. Using serum samples from mice with known influenza exposure history, we showed that our influenza arrays can be used for identification of the infecting influenza subtype. We also showed that sub-lethal infection generates a broad cross-reactive response to H1N1 strains, as well as to H5N1 strains, which are both group 1 influenza strains [21, 22]. Comparing the antibody profiles of individual mice we found extensive heterogeneity in the breadth and magnitude of the response following sub-lethal H1N1 infection - showcasing the ability of the influenza AMs to profile influenza immune-history. We then showed that the arrays can also be used to generate antibody profiles from dried blood spots taken from chickens and that these profiles were highly similar to those measured from the serum of the same chickens. The antibody profiles generated from both the serum and dried blood spots, required minimal sample volumes. Taken together, these data suggest that the influenza antigen microarrays provide a useful alternative to the ELISA assay, especially for profiling antibody responses in small wild animals, as well as from newborns and young children from which very limited sample volumes can be obtained. Using blood spots also does not require any refrigeration in the field, or any centrifugation equipment.

We then optimized the ELISA-on-Chip antigen microarray assay - in which each antigen is spotted in serial dilutions, allowing to calculate an area under the curve (AUC) statistic for each antigen. We found that this was a more robust measure of the antibody levels as compared to measuring antibody levels from a single antigen dilution. While the traditional ELISA assay uses a single concentration of the antigen and requires running each sample in serial dilutions for antibody quantitation when a standard control does not exist, our ELISA-on-Chip assay can estimate antibody levels using a single serum dilution. For example, spotting 16 microarrays on a single 2.4×7.5 cm microarray slide (yielding arrays of 6400 × 6400 μm in size), allows parallel quantification of antibody levels of 40 antigens spotted at 11 dilutions.

We demonstrated that this assay, termed ELISA-on-chip, can be used to quantify binding of mouse monoclonal antibodies, and generates binding curves highly similar to those obtained using the traditional ELISA assay, with overall correlations ranging from 0.71 to 0.95.. Finally, we showed that this assay can be used for rapidly characterizing the specificity and cross-reactivity of a panel of human monoclonal antibodies to a large number of antigens.

Using mouse samples infected with specific influenza strains from the H1N1 and H3N2 subtypes, we showed that the ELISA-on-Chip assay can detect cross-reactive antibody responses within subtype, and that mice infected with H1N1 do not generate an H3N2 response and vice-versa. However, we also found that the assay can capture cross reactivity to other subtypes. In particular, we found that mice infected with H1N1 also generated weaker antibody responses to strains from the H5N1 subtype, which belongs to the same antigenic group [22, 29]. These data suggest that influenza antibody profiles may discriminate between animals exposed to an H1N1 natural infection, vs. an H5N1 natural infection. As such, this may be used as an effective tool for monitoring influenza exposures of wild migrating birds.

The analysis of the human monoclonal antibody profiles generated using our ELISA-on-Chip assay yielded highly related profiles for those obtained for these antibodies in previous studies [14, 15, 26-28]. This showcases the feasibility of using the ELISA-on-Chip assay for rapid mapping of large panels of mAbs. By increasing the initial set of mAbs characterized using this platform, focusing on mAbs with known solved structures and binding properties this platform may in the future be further developed for inferring of the binding footprints of novel mAbs.

Some of the mAbs we tested were more responsive at lower dilutions then others, and this was highly correlated to the idea that different antibodies will have different titers for their activity against their target [30]. In order to overcome this limitation we tested each mAb using a serial dilution and computed the area under the curve (AUC) statistic. Some of the mAbs exhibited moderate binding for their target antigens (i.e. 030-29 2B03), and bound more strongly to other strains, against which they were never previously tested. For example, mAb 051-09 4A03 strongly bound influenza B strains and 217-1A02 had weak binding to H1N1 and H3N2 strains. Some of the mAbs, especially the HA-head specific mAbs had very weak binding to the California 2009 pH1N1 HA (e.g. SFV019 2A02 and 045-09 2B05). This may indicate that these mAbs need to be tested at higher concentrations than the ones used here, and could also be due to improper presentation of the HA head of the California pH1N1 antigen. When weak binding is detected only to a single antigen from a given HA subtype, this may not reflect true cross-reactivity and should be verified with an alternate antibody binding assay.

A limitation of antigen microarrays is batch to batch variability, which has been widely studied across multiple studies [31] [32] [33]. However, in parallel to the availability of multiple standard normalization methods for array data [34], a single batch of slides with 40 antigens per array generates 2240 arrays, which is a sufficiently large number for most studies.

While both the ELISA and ELISA-on-Chip assays are semi-quantitative antibody binding assays, they differ in multiple parameters including: (1) amplification mode colorimetric vs. fluoresentric which in turn also affects their sensitivity and their dynamic range. (2) The ELISA–on-Chip allows multiplexed testing vs. the ELISA assay in which each antigen is tested individually; (3) sample volume requirements - the ELISA-on-Chip assay requires significantly less sample volume as compared to running multiple ELISA assays due to its multiplex nature; (4) Antigen quantity - due to the small spot sizes used on the ELISA-on-ChiP assay, the amount of antigen (and cost) required per sample is significantly lower than the ELISA assay; (5) Testing capacity - the ELISA-on-Chip assay can readily be used to generate antibody profiles for up to 200 samples per day, which would require significantly more time using the traditional ELISA assay even with proper liquid dispensing automation; (6) Fabrication - a clear advantage of the ELISA assay is that plates can be coated manually without the need to sophisticated spotted equipment required for fabricating antigen microarrays. (7) Assay readout - ELISA assays require a plate-reader which is widely available in multiple laboratories. In contrast, scanning arrays requires dedicated laser scanners that are less prevalent. To further illustrate the advantages of the ELISA–on-Chip assay as compared to the traditional ELISA assay, we compared their use for testing 90 samples in two settings: (1) Running each sample at a dilution curve using 4 dilutions-in this setting we compare ELISA using 4 serial dilutions of the sample to ELISA-on-Chip using 4 serial dilutions of the antigen spotted on the AMs (**Table 1** and **Figure 6B**) ; and (2) Running each sample at a single dilution for a single concentration of antigen coated on the ELISA plates or spotted on the AMs (**Table 2**).

**Table 1:** Comparison of ELISA-on-Chip and traditional ELISA for running samples using four serial dilutions. Comparisons assume that 90 individuals are tested for 20 antigens.

|  | AM single antigen | AM 30 antigens | ELISA single antigen | ELISA 30 antigens |
| --- | --- | --- | --- | --- |
| **Number of samples** | 90 | 90 | 90 | 90 |
| **number of slides/plates** | 6<sup>1</sup> | 6 | 3<sup>2</sup> | 90 |
| **antigen quantity**<sup>3</sup> | 2.1 µg<sup>4</sup><br>(triplicate) | - | 74.4 µg<br>(triplicate) | - |
| **antigen concentration**<sup>5</sup> | 65.5-8 ug/ml |  | 4 ug/ml |  |
| **sample volume** | 1 µl | 1 µl | 2 µl (triplicate) | 60 µl |
| **sample dilution (IgG)**<sup>6</sup> | 1:100 |  | 1:100 -1:800 |  |
| **Work time (hours)** | 6 h | 6h | 6h | 120h<sup>7</sup> |

<sup>1</sup> Using microarray slides with 16 arrays per slide each including 20 antigens spotted at 4 dilutions in triplicates.
<sup>2</sup> Using 384-well format ELISA where each sample is tested at 4 dilutions using triplicate wells. 30 individuals can be tested on a single 384 well plate.
<sup>3</sup> Antigen quantity per single antigen assuming triplicate spots (AM) or wells (ELISA)
<sup>4</sup> 2.08 µg will be sufficient to spot 140 microarray slides - sufficient for testing 2100 samples.
<sup>5</sup> Antigens are spotted on the AM at 4 dilutions starting at 65.5 ug/ml at 2-fold dilutions
<sup>6</sup> Minimal dilution used for IgG profiling
<sup>7</sup> Assuming 6 384 well ELISA plates can be run on a single day

**Table 2:** Comparison of ELISA-on-Chip and traditional ELISA for running samples using a single dilution. Comparisons assume that 90 individuals are tested across 40 antigens.

|  | AM single antigen | AM 120 antigens | ELISA single antigen | ELISA 120 antigens |
| --- | --- | --- | --- | --- |
| **Number of samples** | 90 | 90 | 90 | 90 |
| **number of slides/plates** | 6<sup>1</sup> | 6 | 1<sup>2</sup> | 90<sup>3</sup> |
| **antigen quantity**<sup>4</sup> | 0.5 µg (triplicate) | - | 18.6 µg (triplicate) | - |
| **antigen concentration** | 32.5 ug/ml | 32.ug/ml | 4 ug/ml | 4 ug/ml |
| **sample volume** | 1 µl | 1 µl | 1 µl (triplicate) | 108 µl |
| **sample dilution (IgG)**<sup>5</sup> | 1:100 |  | 1:100 |  |
| **Work time (hours)** | 6 h | 6h | 4h | 90h<sup>6</sup> |

<sup>1</sup>Using microarray slides with 16 arrays per slide each including 40 antigens spotted at a single dilution in triplicates.
<sup>2</sup>To test 90 individuals in triplicates we need ¾ of a 384-well plate is required.
<sup>3</sup>Using 384-well format ELISA where each sample is tested using triplicate wells. 90 samples can be run using ¾ of a single plate.
<sup>4</sup>Antigen quantity per single antigen assuming triplicate spots (AM) or wells (ELISA)
<sup>5</sup>Minimal dilution used for IgG profiling
<sup>6</sup>assuming 6 384 well ELISA plates can be run on a single day

While both the ELISA and ELISA-on-Chip assays are semi-quantitative antibody binding assays, they differ in multiple parameters. While the ELISA assay uses an enzyme substrate reaction to produce readable signals, this significantly amplifies the signal allowing increased sensitivity. The AM assay uses fluorescence detection that does not have an enzymatic amplification effect. Therefore, the ELISA assay may have a higher dynamic range across which it may detect antibodies. One possible solution for this issue, which was used here, is the concentration of antigens used for array spotting. Since antigens are spotted in micrometer spots using nanodrops (300-360 pl), antigens can be cost-effectively spotted at much higher concentrations than those used to coat ELISA plates. Nevertheless, our results demonstrate that the ELISA-on-Chip assay produces qualitatively similar results to the traditional ELISA assay. While the AM better distinguishes between the different samples, the ELISA assay has a wider dynamic range due to the significant enzymatic amplification. The high throughput of the ELISA-on-Chip assay, as well as the low volume of biological material required per assay, make the ELISA-on-Chip assay an appropriate alternative antibody binding method for large scale screening. A clear advantage of the ELISA-on-Chip assay over traditional ELISA is the ability to test multiple antigens simultaneously, allowing to rapidly generate antibody binding profiles to hundreds of antigens simultaneously using low volume samples - which is particularly important in studies of newborns and young children.

In summary, here we presented ELISA-on-chip - a novel antigen microarray based ELISA assay and compared it to the traditional ELISA assay. While the ELISA-on-Chip assay cannot be readily performed in any laboratory, and requires access to dedicated laboratory equipment, we demonstrated several antibody profiling applications in which the traditional ELISA assays are not feasible. The ability to perform high-throughput antibody profiling using minimal sample volumes allows to rapidly and cost-effectively screen large datasets, and can be used as a filtering step to identify important samples and antigens that should be further studied using functional antibody assays.

## Data Availability

All row data and code used in the analysis are available in the Hertz Lab website: https://www.hertz-lab.org/

---

**Supplementary Figure 1:**
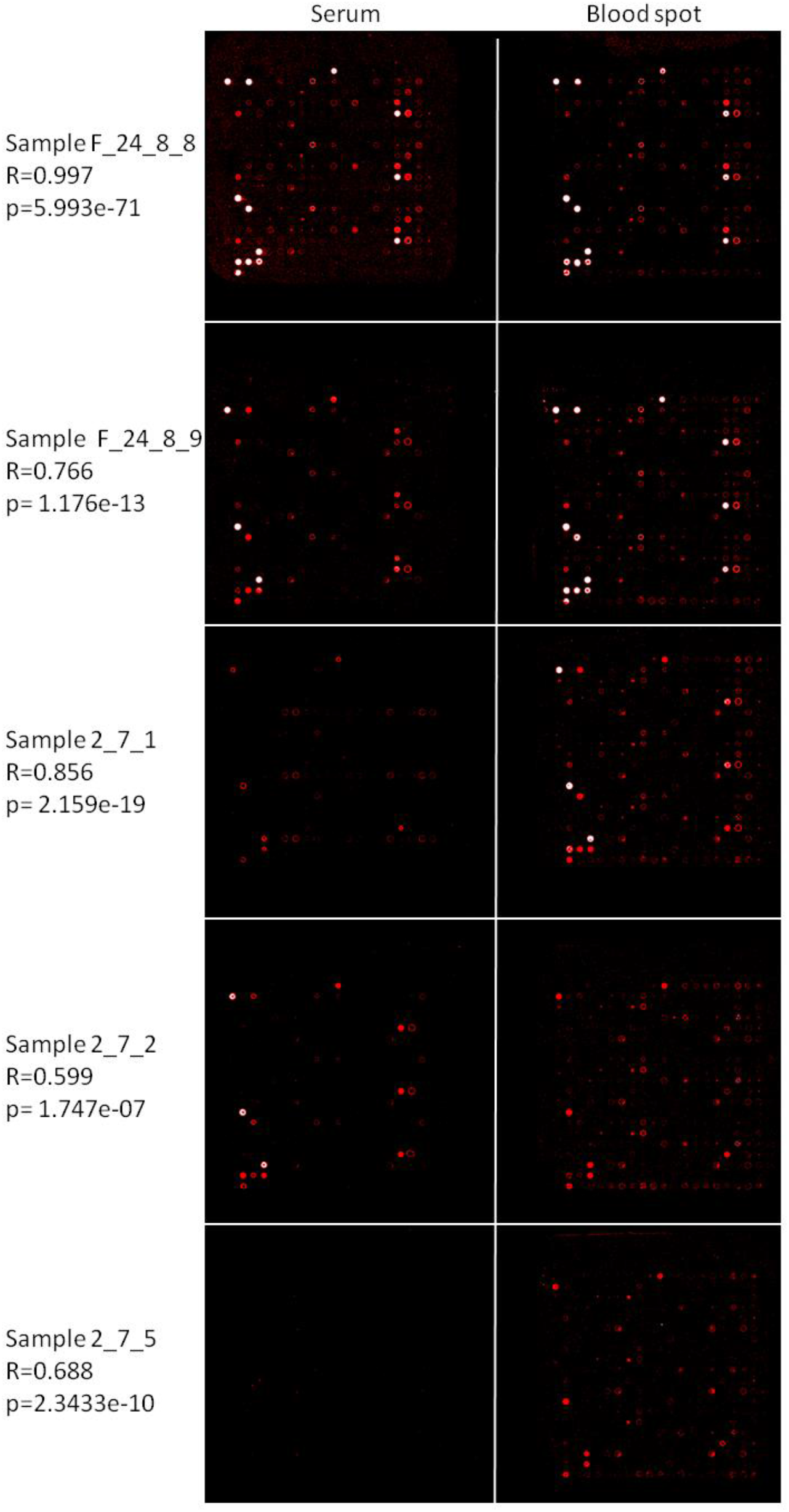
Examples for chicken serum-blood spot AM pairs with low correlations. Serum and dry blood spots were collected from the same blood samples of chickens vaccinated with the H9N2 vaccine, and the anti-influenza IgY repertoires were measured using AMs spotted with influenza recombinant proteins at a single concentration. Pearson correlations (R and p values) were computed between AM results for the serum sample (diluted 1:4000; left panels) and the dry blood spot sample (reconstituted in 2 ml buffer and diluted 1:20; right panels) that were collected from the same blood sample. The upper row is an example for a serum-blood spot pair from the chicken with an excellent correlation. The lower 4 rows display the AM pairs for 4/8 sample pairs with low correlations, as an example. In all these pairs the anti-influenza IgY level was low, and as a result, staining of many influenza antigens was not observed with the serum samples, although it was observed using the blood spot samples, which are more sensitive. The lack of antibody detection is the serum responsible for the low Pearson correlations in these sample pairs.

**Supplementary Table 1:**
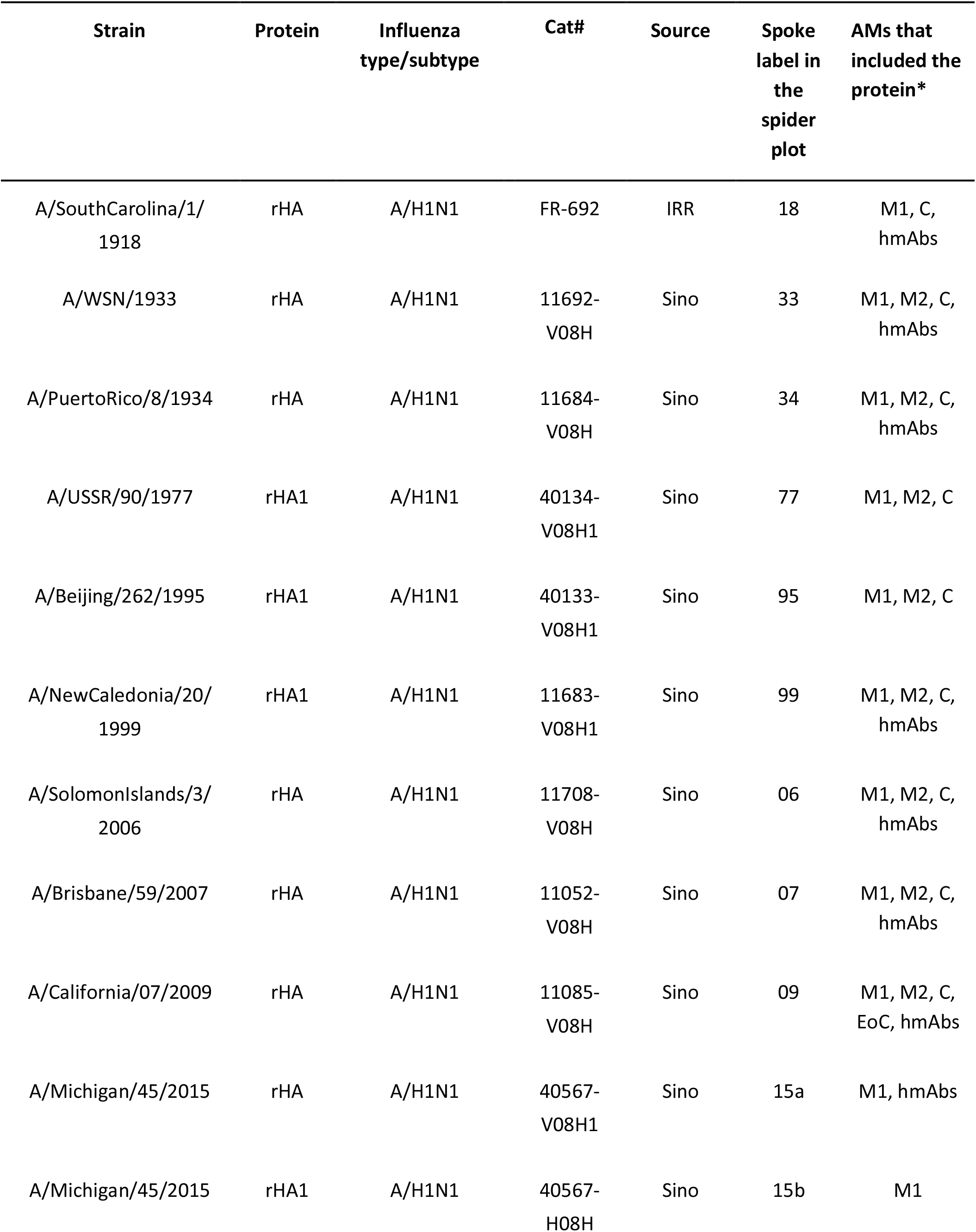
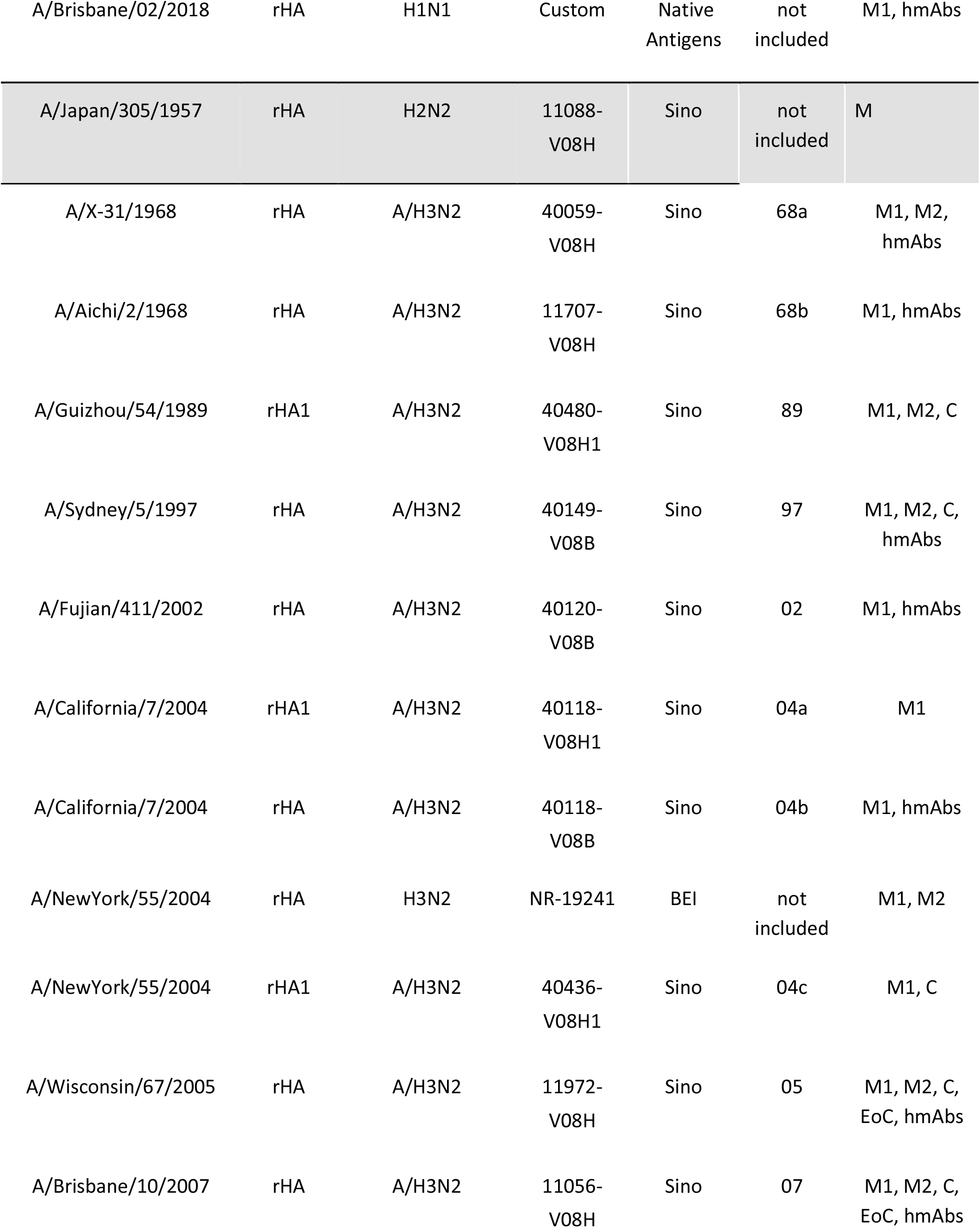
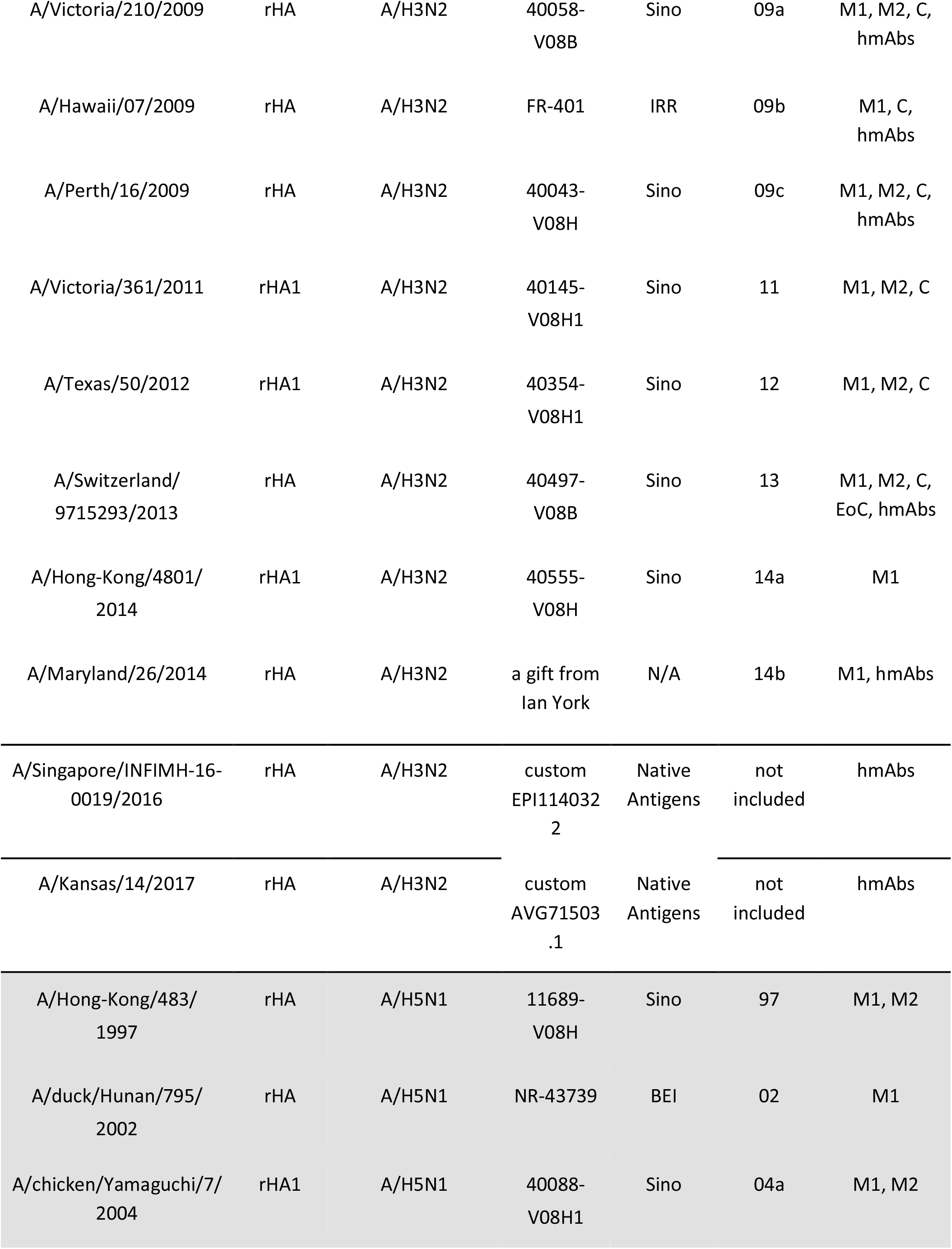
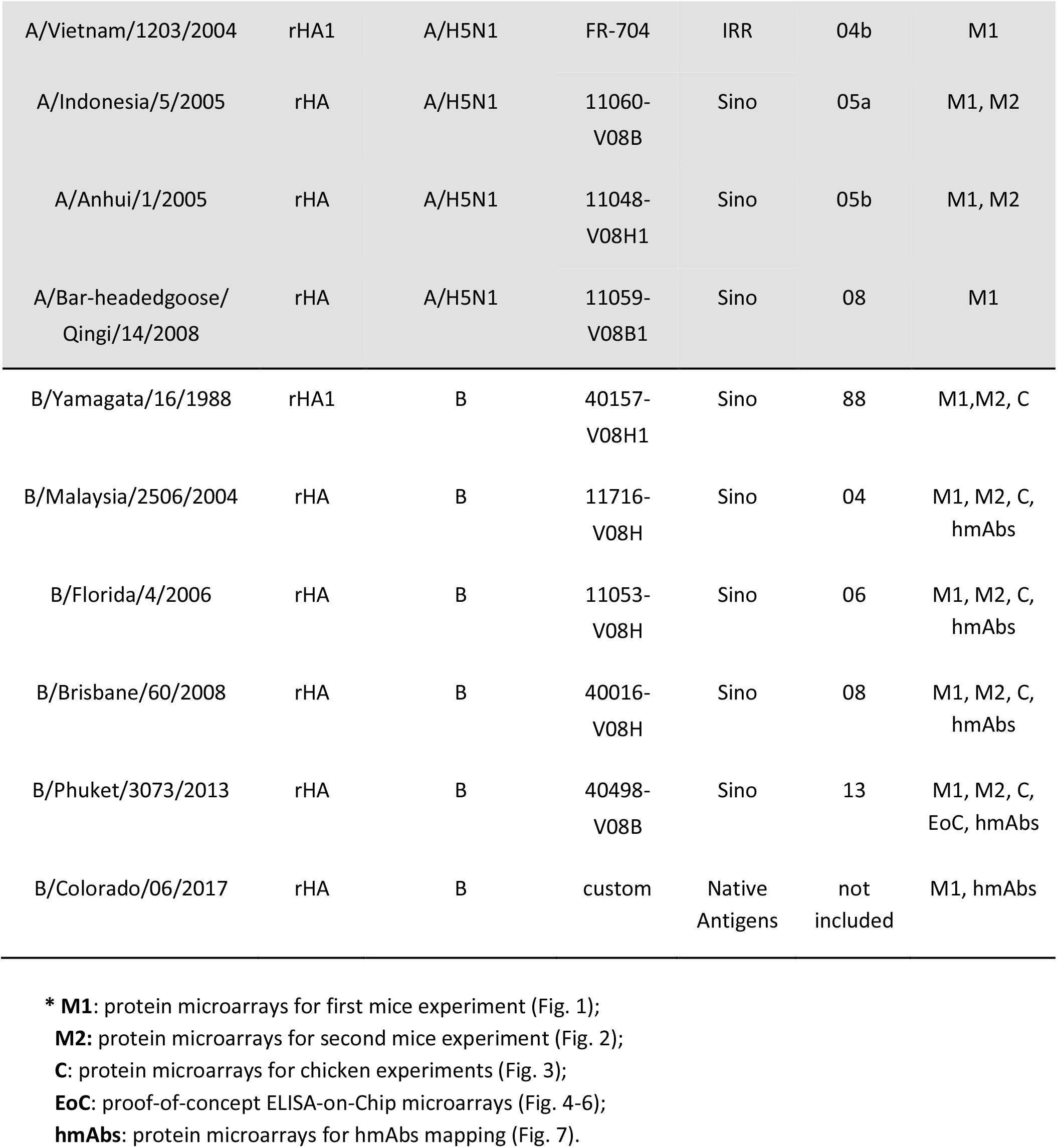
Recombinant hemagglutinin proteins from human and avian influenza strains, spotted on the protein antigen microarray chips.

**Supplementary Table 2:** Spearman correlation coefficients comparing the traditional ELISA assay inter-correlations. Correlations were computed using the AUC values of 10 individuals for each antigen separately.

|  | **Brisbane**<br>**ELISA** | **California**<br>**ELISA** | **Wisconsin**<br>**ELISA** | **Shanghai**<br>**ELISA** |
| --- | --- | --- | --- | --- |
| **Brisbane**<br>**ELISA** | 1 | 0.67 | 0.89 | 0.50 |
| **California**<br>**ELISA** | 0.67 | 1 | 0.83 | 0.41 |
| **Wisconsin**<br>**ELISA** | 0.89 | 0.83 | 1 | 0.64 |
| **Shanghai**<br>**ELISA** | 0.50 | 0.41 | 0.64 | 1 |

**Supplementary Table 3:** Spearman correlation coefficients comparing the ELISA-on-Chip assay inter-correlations. Correlations were computed using the AUC values of 10 individuals for each antigen separately for samples diluted 1:3200.

|  | **Brisbane**<br>**AM** | **California**<br>**AM** | **Wisconsin**<br>**AM** | **Shanghai**<br>**AM** |
| --- | --- | --- | --- | --- |
| **Brisbane**<br>**AM** | 1 | 0.64 | 0.42 | 0.025 |
| **California**<br>**AM** | 0.64 | 1 | 0.81 | 0.50 |
| **Wisconsin**<br>**AM** | 0.42 | 0.81 | 1 | 0.58 |
| **Shanghai**<br>**AM** | 0.025 | 0.50 | 0.58 | 1 |

**Supplementary Table 4:** Human monoclonal antibodies used in this study.

| mAb | Binding reference | Antigen | Epitope | Function | Reference |
| --- | --- | --- | --- | --- | --- |
| SFV019-2A02 | H1 | Head | Ca1 | HAI+ | 35, 36 |
| SFV015-2F04 | H1 | Head | Lateral Patch | HAI+ | 35, 36 |
| 045-09 2B05 | H1 | Head | Lateral Patch | HAI+ | 15, 36 |
| 030-09 2B03 | H1 | Stalk | BN | Neut+ | 15, 36 |
| CR-9114 | H1, H3, B | Stalk | BN | Neut+ | 26, 28, 37 |
| FI6 | H1, H3 | Stalk | BN | Neut+ | 26 ,28 ,37 |
| 047-09 1G05 | H1 | Stalk | Stalk | Neut- | 15 |
| 030-09 1E05 | H1, H3, B | Stalk | Stalk | Neut- | 15 |

## Author contributions

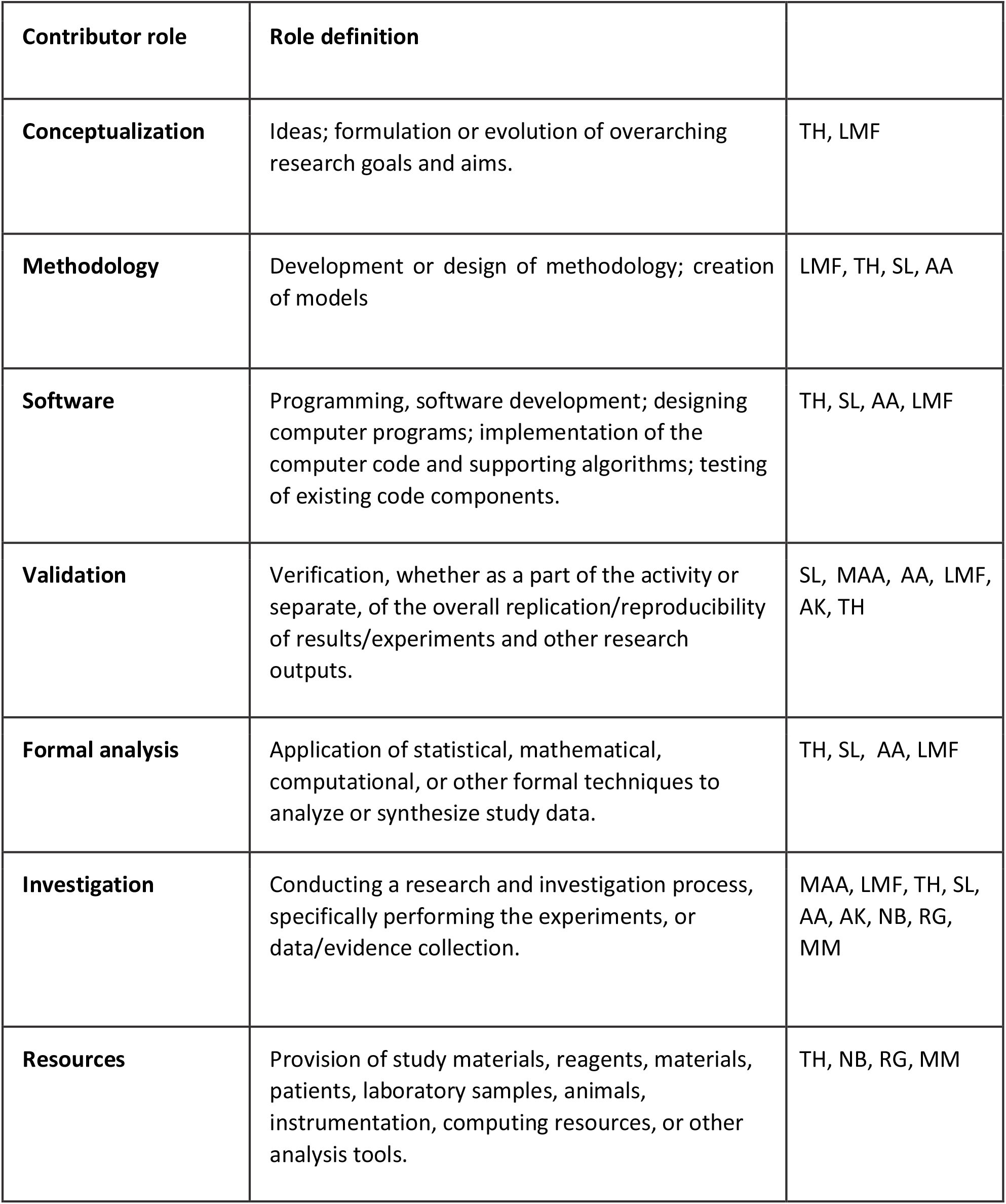
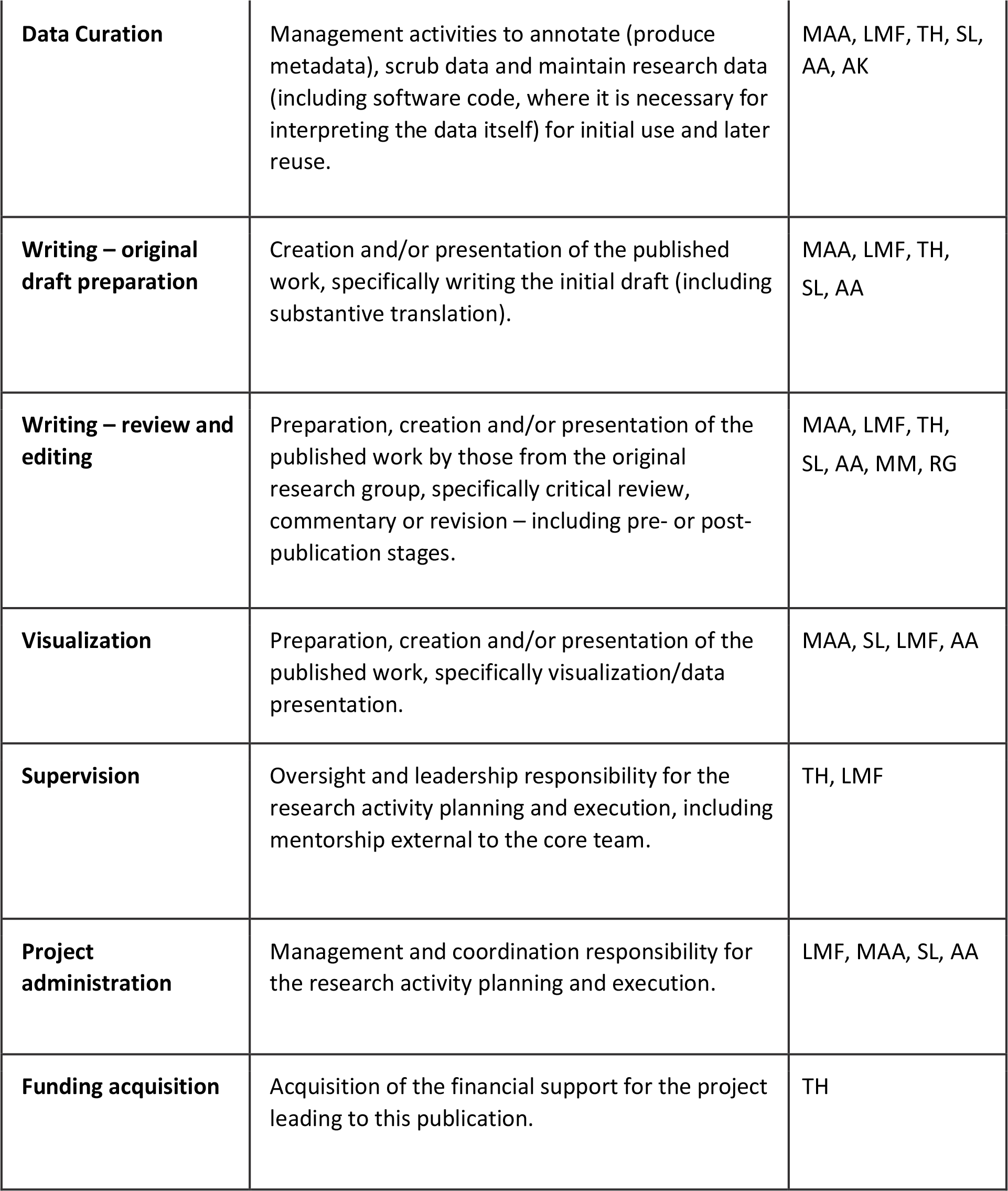

## Reference

1. Engvall, E. and P. Perlmann, Enzyme-linked immunosorbent assay (ELISA). Quantitative assay of immunoglobulin G. Immunochemistry, 1971. 8(9): p. 871–4.

2. Engvall, E., K. Jonsson, and P. Perlmann, Enzyme-linked immunosorbent assay. II. Quantitative assay of protein antigen, immunoglobulin G, by means of enzyme-labelled antigen and antibody-coated tubes. Biochim Biophys Acta, 1971. 251(3): p. 427–34.

3. Aydin, S., A short history, principles, and types of ELISA, and our laboratory experience with peptide/protein analyses using ELISA. Peptides, 2015. 72: p. 4–15.

4. Mikulskis, A., et al., Solution ELISA as a platform of choice for development of robust, drug tolerant immunogenicity assays in support of drug development. J Immunol Methods, 2011. 365(1-2): p. 38–49.

5. Lai, S., et al., Design of a compact disk-like microfluidic platform for enzyme-linked immunosorbent assay. Anal Chem, 2004. 76(7): p. 1832–7.

6. Ng, S., et al., Novel correlates of protection against pandemic H1N1 influenza A virus infection. Nat Med, 2019. 25(6): p. 962–967.

7. Price, J.V., et al., Characterization of influenza vaccine immunogenicity using influenza antigen microarrays. PLoS One, 2013. 8(5): p. e64555.

8. Kunnath-Velayudhan, S., et al., Proteome-scale antibody responses and outcome of Mycobacterium tuberculosis infection in nonhuman primates and in tuberculosis patients. J Infect Dis, 2012. 206(5): p. 697–705.

9. Keating, R., et al., The kinase mTOR modulates the antibody response to provide cross-protective immunity to lethal infection with influenza virus. Nat Immunol, 2013. 14(12): p. 1266–76.

10. Zhong, L., et al., Using protein microarray as a diagnostic assay for non-small cell lung cancer. Am J Respir Crit Care Med, 2005. 172(10): p. 1308–14.

11. Quintana, F.J., et al., Antigen microarrays identify unique serum autoantibody signatures in clinical and pathologic subtypes of multiple sclerosis. Proc Natl Acad Sci U S A, 2008. 105(48): p. 18889–94.

12. Merbl, Y., et al., Newborn humans manifest autoantibodies to defined self molecules detected by antigen microarray informatics. J Clin Invest, 2007. 117(3): p. 712–8.

13. Gazit, R., et al., Lethal influenza infection in the absence of the natural killer cell receptor gene Ncr1. Nat Immunol, 2006. 7(5): p. 517–23.

14. Guthmiller, J.J., et al., An Egg-Derived Sulfated N-Acetyllactosamine Glycan Is an Antigenic Decoy of Influenza Virus Vaccines. mBio, 2021. 12(3): p. e0083821.

15. Andrews, S.F., et al., Immune history profoundly affects broadly protective B cell responses to influenza. Sci Transl Med, 2015. 7(316): p. 316ra192.

16. Nachbagauer, R., et al., Broadly Reactive Human Monoclonal Antibodies Elicited following Pandemic H1N1 Influenza Virus Exposure Protect Mice against Highly Pathogenic H5N1 Challenge. J Virol, 2018. 92(16).

17. Mahallawi, W.H., et al., Infection with 2009 H1N1 influenza virus primes for immunological memory in human nose-associated lymphoid tissue, offering cross-reactive immunity to H1N1 and avian H5N1 viruses. J Virol, 2013. 87(10): p. 5331–9.

18. Jegaskanda, S., et al., Cross-reactive influenza-specific antibody-dependent cellular cytotoxicity antibodies in the absence of neutralizing antibodies. J Immunol, 2013. 190(4): p. 1837–48.

19. Carreno, J.M., et al., H1 Hemagglutinin Priming Provides Long-Lasting Heterosubtypic Immunity against H5N1 Challenge in the Mouse Model. mBio, 2020. 11(6).

20. Ahmed, M.S., et al., Cross-reactive immunity against influenza viruses in children and adults following 2009 pandemic H1N1 infection. Antiviral Res, 2015. 114: p. 106–12.

21. Gostic, K.M., et al., Potent protection against H5N1 and H7N9 influenza via childhood hemagglutinin imprinting. Science, 2016. 354(6313): p. 722–726.

22. Arevalo, C.P., et al., Original antigenic sin priming of influenza virus hemagglutinin stalk antibodies. Proc Natl Acad Sci U S A, 2020. 117(29): p. 17221–17227.

23. Zost, S.J., et al., Rapid isolation and profiling of a diverse panel of human monoclonal antibodies targeting the SARS-CoV-2 spike protein. Nat Med, 2020. 26(9): p. 1422–1427.

24. Rogers, T.F., et al., Isolation of potent SARS-CoV-2 neutralizing antibodies and protection from disease in a small animal model. Science, 2020. 369(6506): p. 956–963.

25. Pedrioli, A. and A. Oxenius, Single B cell technologies for monoclonal antibody discovery. Trends Immunol, 2021. 42(12): p. 1143–1158.

26. Ekiert, D.C., et al., A highly conserved neutralizing epitope on group 2 influenza A viruses. Science, 2011. 333(6044): p. 843–50.

27. Dreyfus, C., et al., Highly conserved protective epitopes on influenza B viruses. Science, 2012. 337(6100): p. 1343–8.

28. Corti, D., et al., A neutralizing antibody selected from plasma cells that binds to group 1 and group 2 influenza A hemagglutinins. Science, 2011. 333(6044): p. 850–6.

29. Knight, M., et al., Imprinting, immunodominance, and other impediments to generating broad influenza immunity. Immunol Rev, 2020. 296(1): p. 191–204.

30. Barrette, R.W., J. Urbonas, and L.K. Silbart, Quantifying specific antibody concentrations by enzyme-linked immunosorbent assay using slope correction. Clin Vaccine Immunol, 2006. 13(7): p. 802–5.

31. Zhu, H., et al., Autoantigen Microarray for High-throughput Autoantibody Profiling in Systemic Lupus Erythematosus. Genomics Proteomics Bioinformatics, 2015. 13(4): p. 210–8.

32. Ayoglu, B., J.M. Schwenk, and P. Nilsson, Antigen arrays for profiling autoantibody repertoires. Bioanalysis, 2016. 8(10): p. 1105–26.

33. Brezina, S., et al., Immune-Signatures for Lung Cancer Diagnostics: Evaluation of Protein Microarray Data Normalization Strategies. Microarrays (Basel), 2015. 4(2): p. 162–87.

34. Hamelinck, D., et al., Optimized normalization for antibody microarrays and application to serum-protein profiling. Mol Cell Proteomics, 2005. 4(6): p. 773–84.

35. Li, G.M., et al., Pandemic H1N1 influenza vaccine induces a recall response in humans that favors broadly cross-reactive memory B cells. Proc Natl Acad Sci U S A, 2012. 109(23): p. 9047–52.

36. Guthmiller, J.J., et al., First exposure to the pandemic H1N1 virus induced broadly neutralizing antibodies targeting hemagglutinin head epitopes. Sci Transl Med, 2021. 13(596).

37. Guthmiller, J.J., et al., Broadly neutralizing antibodies target a haemagglutinin anchor epitope. Nature, 2022. 602(7896): p. 314–320.

